# A non-invasive liquid biopsy resolves the diagnostic blind spot in chronic kidney disease

**DOI:** 10.64898/2026.06.09.26355142

**Authors:** Tatsuya Nishimura, Yutaka Harita, Yosuke Hirakawa, Keiichi Takizawa, Jun Fujishiro, Sumito Ogawa, Yuko Kajiho, Shoichiro Kanda, Reiko Kushima, Tae Omori, Yuko Hamasaki, Yoshimitsu Gotoh, Kenichiro Miura, Naoya Fujita, Takayuki Okamoto, Masataka Hisano, Masaomi Nangaku, Motohiro Kato

## Abstract

Chronic kidney disease is a major global health burden, and its early detection is critical for delaying progression to kidney failure using recently developed targeted therapies. However, current diagnostic screening relies heavily on blood markers that are confounded by muscle mass, and on urine tests that frequently miss structural damage occurring without protein leakage. This creates a critical diagnostic blind spot that hinders timely intervention. Here we show a non-invasive liquid biopsy platform that quantifies a specific protein marker, MUC1, on urinary extracellular vesicles to accurately assess renal parenchymal integrity. By bypassing the systemic metabolic noise of traditional blood tests, our assay provides a remarkably stable, person-specific functional signature. Following extensive validation across diverse cohorts, our longitudinal analysis demonstrated that the discrepancy between this novel urine-based readout and standard blood tests unmasks hidden renal vulnerability, successfully predicting rapid functional decline. By comprehensively evaluating both tubular and glomerular integrity from a single spot urine sample, these findings establish a completely non-invasive, highly scalable prescreening tool that resolves the diagnostic blind spot, enabling broader early detection strategies and ushering in a new era of proactive risk management.

## Introduction

Chronic kidney disease (CKD) is a major global health burden, affecting more than 700 million individuals and contributing substantially to cardiovascular morbidity and mortality^1,2^. While the advent of sodium–glucose cotransporter 2 inhibitors (SGLT2i) provides a pivotal opportunity to delay progression to end-stage kidney disease when introduced early^3–8^, their full potential remains unrealized because current screening strategies frequently fail to identify early renal decline^9–11^.

At present, CKD is diagnosed and staged using a combination of estimated glomerular filtration rate (eGFR) and albuminuria/proteinuria. Although this framework is foundational, it contains a critical “CKD blind spot,” in which substantial renal impairment remains undetected^1,10,12,13^. The most common eGFR formulas are based on serum creatinine (sCre), which is a byproduct of muscle metabolism and thus heavily influenced by age, sex, and muscle mass^2,14,15^. While cystatin C (CysC)-based eGFR provides an alternative, its clinical reliability can be confounded by factors such as inflammation, thyroid status, or steroid therapy^8^. Recent large-scale meta-analyses have highlighted that discordance between different filtration markers, such as the gap between sCre - and CysC -based eGFR (eGFRcr and eGFRcysC, respectively) is a potent predictor of adverse clinical outcomes, including kidney failure and cardiovascular mortality, suggesting that such “gaps” unmask hidden renal vulnerability^16^. Furthermore, up to two-thirds of patients with moderate-to-severe CKD lack significant proteinuria, allowing non-proteinuric phenotypes to escape routine early detection until irreversible damage has accumulated^17–20^.

To address this unmet need, a urinary extracellular vesicle (uEV)-based “liquid biopsy” has been proposed as a means to capture segment-specific information from the nephron^21–26^. uEVs are nanoscale vesicles released by virtually all nephron segments and carry proteins and transcripts that reflect the state of the renal microenvironment^25^. In earlier work in a pediatric cohort, proteomic profiling identified Mucin-1 (MUC1) on uEVs as a promising biomarker, with decreased MUC1 levels in uEVs closely associated with declining renal function^27,28^. However, translating this biological insight into a clinical tool requires demonstrating generalizability across adult etiologies, establishing a high-throughput assay, and confirming large-scale analytical stability.

Here, we present a standardized, non-invasive liquid biopsy platform targeting MUC1-positive uEVs. Evaluated in a diverse multicenter cohort of 860 individuals, MUC1 in uEVs provides a sensitive readout of renal functional decline even when traditional markers remain uninformative. By integrating this marker with proteinuria, we demonstrate refined stratification for SGLT2i therapy and introduce the “eGFR-Gap” to identify high-risk individuals, supporting a shift in CKD management toward proactive, biology-based intervention

## Results

### MUC1 in uEVs Captures Renal Functional Decline across a Diverse Multicenter Cohort

To quantify MUC1 within uEVs, we utilized two distinct capture platforms: Tim4-coated and CD9-coated ELISA plates. Two MUC1-detection configurations—MUC1_Tim4_ (Tim4-capture/MUC1Ab-detection) and MUC1_CD9_ (CD9Ab-capture/MUC1Ab-detection)—were used. To ensure robust comparability across the multiple assays and cohorts, measured MUC1 levels were standardized into Reference Units (RU), where 100 RU was defined by the baseline level of healthy young adults (aged < 40 years, eGFRcr ≥ 90 mL/min/1.73 m²). We evaluated the diagnostic performance of these platforms using 1,188 urine samples from a diverse multicenter cohort of 860 individuals, stratified into a Discovery Cohort (n = 471)(Figure 1a), a longitudinal Hospital Cohort (Validation-HC, n = 192) (Figure 2a), and a community Biobank Cohort (Validation-BC, n = 197) (Extended Figure 2a)(Extended Table 1).

**Figure 1.**
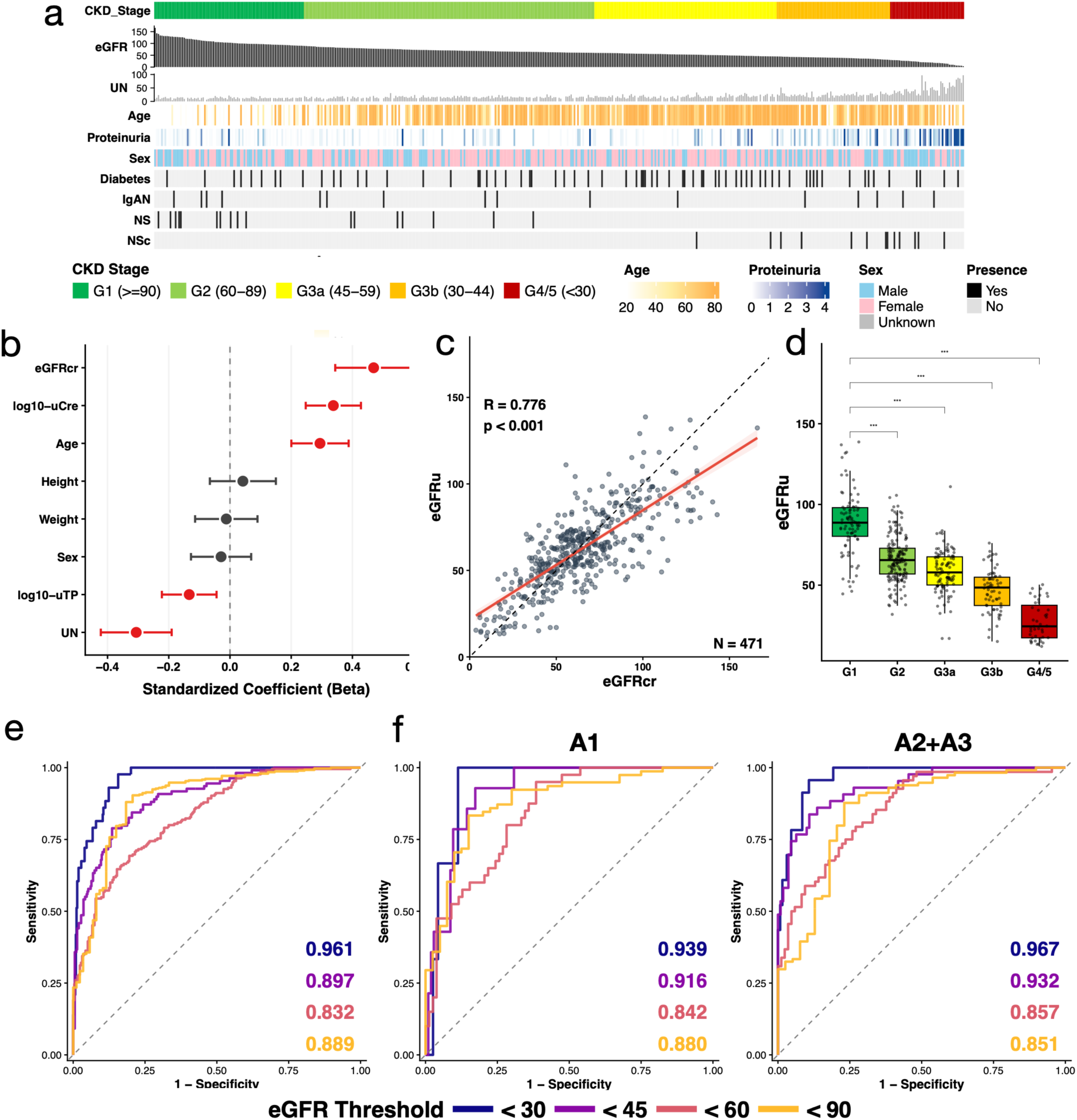
Development and validation of the uEV-based eGFR model (eGFRu). a, Clinical and molecular landscape of the discovery cohort (n = 471). b, Standardized regression coefficients (β) for predictors of urinary MUC1_CD9_ levels, including eGFRcr, log-transformed urinary creatinine, age, height, sex, weight, log-transformed urinary total protein (uTP), and urea nitrogen (UN). c, Scatter plot of eGFRu versus eGFRcr in the discovery cohort. The solid red line represents the linear regression line, and the shaded area indicates the 95% confidence interval. The dashed black line represents the identity line (y=x). The correlation between eGFRcr and eGFRu was assessed using Pearson’s correlation coefficient. d, The box-and-whisker plots represent the distribution of eGFRu, categorized by clinical CKD stages (G1 to G4/5) based on reference eGFRcr. Statistical significance was determined using the Wilcoxon rank-sum test, comparing each stage to the G1 group (\*\*\**p* < 0.001). e, ROC curves of eGFRu for identifying stages of renal impairment defined by eGFRcr thresholds (< 30, 45, 60, and 90 mL/min/1.73 m²). f, ROC curves of eGFRu for identifying renal impairment stratified by normoalbuminuria (left; ACR < 30 mg/gCr, stage A1) and clinically significant proteinuria (right; ACR ≥ 30 mg/gCr, stages A2–A3). Scatter plots show individual data points with linear regression lines and 95% confidence intervals (shaded areas).

**Figure 2.**
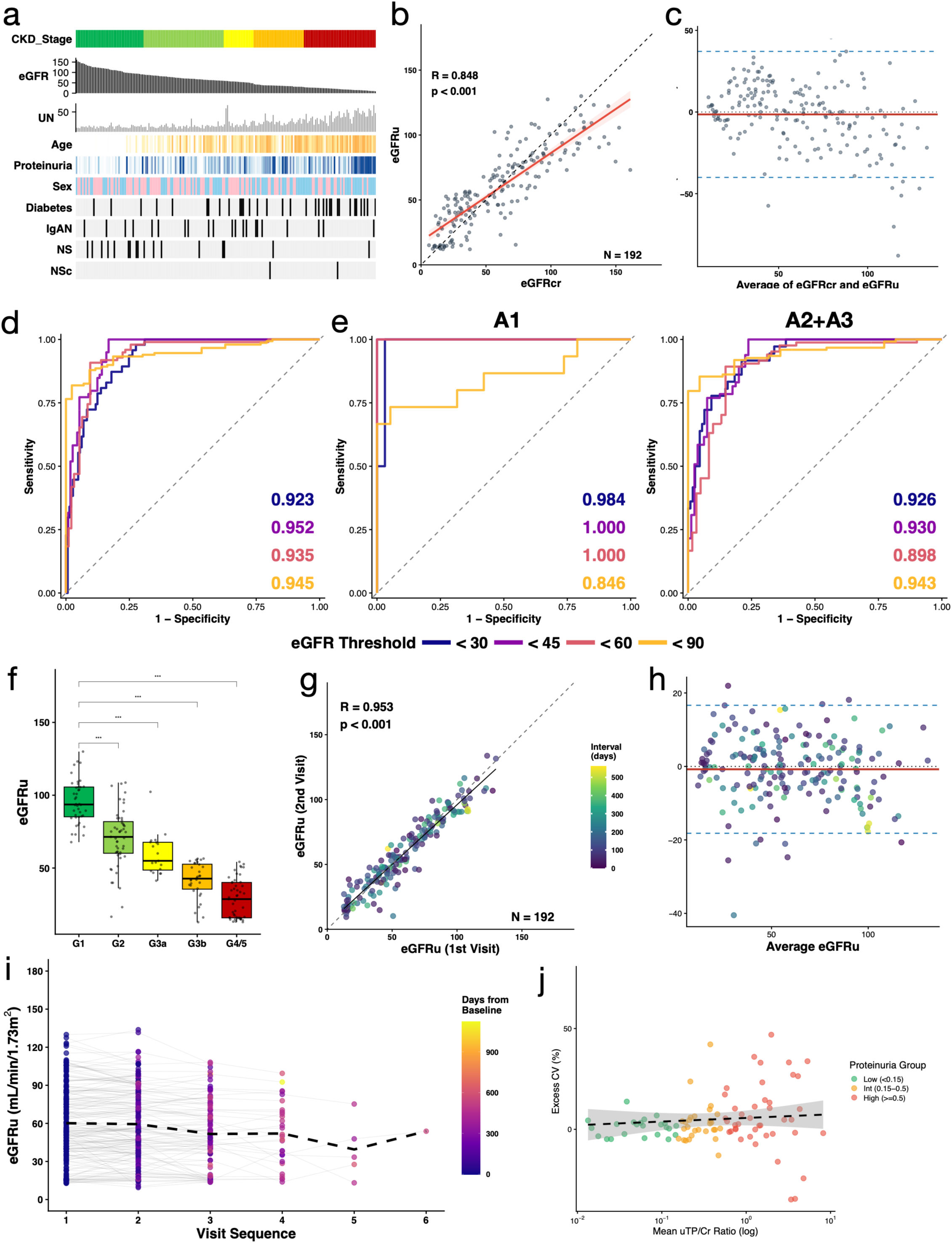
Validation of eGFRu in a prospective cohort and assessment of longitudinal stability. a, Clinical and molecular landscape of the prospective hospital-based validation cohort (Validation-HC; n = 192). b, Correlation between eGFRu and eGFRcr at baseline. c, Bland–Altman plot of agreement between eGFRu and eGFRcr. d, ROC curves of eGFRu for identifying stages of renal impairment defined by eGFRcr thresholds (< 30, 45, 60, and 90 mL/min/1.73 m²). e, ROC curves of eGFRu for identifying renal impairment stratified by normoalbuminuria (left; ACR < 30 mg/gCr, stage A1) and clinically significant proteinuria (right; ACR ≥ 30 mg/gCr, stages A2–A3). f, The box-and-whisker plots represent the distribution of eGFRu, categorized by clinical CKD stages (G1 to G4/5) based on reference eGFRcr. Statistical significance was determined using the Wilcoxon rank-sum test, comparing each stage to the G1 group (\*\*\**p* < 0.001). g, h, Test–retest reliability of eGFRu: scatter plot of two consecutive measurements (g) and the corresponding Bland–Altman plot (h). i, Longitudinal tracking of eGFRu across multiple follow-up visits; data points are colour-coded by time from baseline (days). j, Relationship between excess coefficient of variation (CV) of eGFRu and baseline urinary protein load. Scatter plots include linear regression lines with 95% confidence intervals (shaded areas). The correlation between eGFRcr and eGFRu was assessed using Pearson’s correlation coefficient (b, g).

In the Discovery Cohort, both raw and urinary creatinine (uCre)-normalized MUC1_Tim4_ and MUC1_CD9_ levels demonstrated significant positive correlations with eGFRcr, regardless of normalization for uCre (Extended Figure 1a-d). Normalization by uCre yielded moderate but highly significant correlations, yielding ρ = 0.512(*p* = 6.69 × 10^−33^) for MUC1_Tim4_/uCre and ρ = 0.537 (*p* = 1.49 × 10^−36^) for MUC1_CD9_/uCre. While both platforms demonstrated constant associations with global renal function (Extended Figure 1a-d) and high diagnostic accuracy, particularly for identifying advanced CKD stages (Extended Figure 1e-h), the CD9-capture platform provided superior predictive accuracy for eGFRcr, yielding a higher Adjusted *R*² (0.534 vs. 0.432) and a lower Akaike Information Criterion (AIC) (382.4 vs. 476.1) than the Tim4 platform. Consequently, the CD9-capture platform was selected as the primary assay for all subsequent main analyses.

To highlight the specific advantage of the uEV-based platform, we compared these results with conventional soluble urinary biomarkers, namely free urinary free MUC1 (ufMUC1) (Extended Figure 1i, k, m, o) and NGAL (uNGAL) ^29–31^(Extended Figure 1j, l, n, p). In stark contrast to the uEV-bound markers, both ufMUC1 and uNGAL exhibited substantially weaker correlations with eGFRcr, regardless of uCre normalization (e.g., ρ = 0.319 for ufMUC1/uCre, and ρ = -0.433 for uNGAL/uCre)(Extended Figure 1i-l). Furthermore, their diagnostic performance for identifying renal impairment was notably inferior, with AUC values consistently plateauing between 0.62 and 0.76 across all eGFRcr thresholds (Extended Figure 1m-p). These comparative data clearly demonstrate that MUC1 must be evaluated within the context of extracellular vesicles to achieve high-precision functional estimation.

### Determinants of uEVs-MUC1

To elucidate the independent predictors of MUC1_CD9_ levels and establish a statistical foundation for constructing an eGFR estimation model, we performed standardized multiple regression analysis in the Discovery Cohort (Figure 1b, Extended Table 2).

The model evaluated the influence of eight potential factors: Sex, Age (years), height (BH) (cm), weight (BW) (kg), eGFRcr (mL/min/1.73 m²), Urea Nitrogen (UN) (mg/dL), urinary total protein (uTP) (mg/dL), and uCre (mg/dL). For the MUC1_CD9_ platform, the multivariate model demonstrated a strong explanatory power (Adjusted *R*^2^ 0.751, *p* < 0.001). eGFRcr was identified as a dominant independent positive predictor. Log-transformed urinary creatinine and age also emerged as a significant positive driver (Figure 1b, Extended Table 2). Conversely, UN exhibited a significant negative association, and uTP exhibited a weak but significant negative association. Physical parameters, including height and weight, did not significantly influence urinary MUC1_CD9_ levels, suggesting that the signal serves as a robust reporter of renal functional status, largely independent of external physical habitus.

### Establishment and Calibration of a Stable, uEV-Based Functional Score

We developed a non-invasive predictive model for eGFRcr using a multiplicative power-function structure. Before constructing the multivariable model, we evaluated the direct mathematical relationship between eGFRcr and MUC1_CD9_ using a power-function model across all three independent cohorts. Strikingly, the exponent for MUC1_CD9_ consistently converged to approximately 0.25 (0.223 in the Discovery cohort, 0.233 in the Validation-HC, and 0.284 in the Validation-BC), while maintaining a robust explanatory power in all settings (*R*² > 0.50). This consistent mathematical relationship laid a strong foundation for optimizing the unified predictive formula. Next, a 4-variable model (Age, MUC1_CD9_, uCre, Sex) was constructed (Extended Table 3). We identified a near-perfect mathematical symmetry in the coefficients for log_10_(MUC1_CD9_) (+0.256) and log_10_(uCre) (-0.252), indicating that these variables function most effectively as a ratio (Extended Table 3). Notably, Sex was not a significant predictor in this model (*p* = 0.985). Based on this result and the observed mathematical symmetry between MUC1_CD9_ and uCre, we simplified the structure into a 2-variable ratio-based model (Age and MUC1_CD9_/uCre ratio). This ratio-based model exhibited a lower AIC compared to the initial model (272.30 vs. 274.18), with an improvement of 1.88 favoring the simpler structure.

To further streamline the clinical application, we evaluated the incremental benefit of adding Body Surface Area (BSA), which showed no significant improvement (*p* = 0.395). While uTP exhibited a statistically significant association in a subset analysis (*p* < 0.001), its contribution was considered marginal relative to the dominant effects of the core variables. Following the principle of parsimony and clinical accessibility, we finalized the sex-independent 2-variable formula:

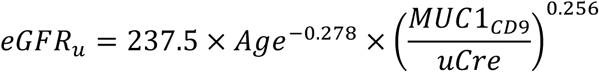

### Discriminative Performance and Clinical Utility in the Discovery Cohort

In the Discovery Cohort, the optimized eGFRu achieved a robust correlation with eGFRcr (*r* = 0.776) (Figure 1c), and exhibited a significant downward trend consistent with increasing CKD severity (Figure 1d). Furthermore, ROC analysis showed excellent accuracy in identifying significant renal impairment, with AUCs of 0.961 (eGFRcr < 30) and 0.897 (eGFRcr < 45) (Figure 1e). The diagnostic performance was strikingly superior in the younger, working-age population (< 60 years). In this subgroup, the model achieved an exceptional AUC of 0.932 and a sensitivity of 81.6% for detecting early functional decline (eGFRcr < 60 mL/min/1.73 m²) (Table 1, Supplemental Table 1), effectively serving as a powerful early screening tool. Furthermore, MUC1_CD9_ maintained high discriminative power even in the normoalbuminuric (A1) group (Figure 1f), confirming its ability to resolve the “CKD blind spot” by capturing intrinsic parenchymal loss independently of glomerular leakage.

**Table 1.** Diagnostic accuracy of eGFRu in the overall population and in adults aged < 60 years across three cohorts.

| Cohort / Population | n | AUC (<60) | Sensitivity | Specificity |
| --- | --- | --- | --- | --- |
| Discovery<br>(eGFR<sub>cr</sub>) | 471 | 0.832 | 69.3% | 80.5% |
| Age < 60 | 183 | 0.932 | 81.6% | 94.0% |
| Validation_HC<br>(eGFR<sub>cr</sub>) | 192 | 0.935 | 90.8% | 90.4% |
| Age < 60 | 131 | 0.937 | 95.7% | 82.1% |
| Validation_BC<br>(eGFR<sub>cysC</sub>) | 197 | 0.925 | 92.1% | 78.0% |
| Age < 60 | 127 | 0.971 | 86.7% | 97.3% |

Sensitivity and specificity are shown as percentages. AUC, area under the receiver operating characteristic curve; eGFR<sub>cr</sub>, creatinine-based estimated glomerular filtration rate; eGFR<sub>cysC</sub>, cystatin C-based estimated glomerular filtration rate.

### Cystatin C Alignment in Independent Validation in the Biobank Cohort

To confirm the generalizability and robustness of our findings, we evaluated the eGFRu formula across two independent validation cohorts. First, we performed a validation study using the community-based Biobank Cohort (Validation-BC) (n = 197) (Extended Figure 2a), where Cystatin C (CysC) measurements were available. Despite decadal cryopreservation (collected from 2011 onwards), the eGFRu formula demonstrated a robust correlation with eGFRcr (*r* = 0.676) (Extended Figure 2b, c).

In this cohort, while the eGFRu formula identified eGFRcr-defined impairment with moderate accuracy (Extended Figure 2d), it achieved a strikingly higher accuracy for identifying CysC-defined impairment (eGFRcysC < 60) (AUC = 0.925) (Extended Figure 2e, Table 1), which further improved to an exceptional AUC of 0.971 in the younger demographic (age < 60) (Supplemental Table 1). This remarkable alignment with a muscle mass-independent filtration marker confirms that eGFRu successfully overcomes the inherent biases of creatinine-based estimation.

### Mathematical Resilience to Muscle Mass Bias

Mathematical modeling confirmed that normalizing the MUC1_CD9_ by uCre internalizes gender-dependent muscular factors. The age-dependent exponent in our formula (−0.278) showed remarkable mathematical symmetry with those of both the established JSN (−0.287) and MDRD (−0.203) equations^32,33^, aligning closely with the mathematical structure of clinical filtration markers. Adjusting female sCre to a male baseline caused data from all cohorts to converge onto a single, gender-invariant trend line. Notably, this convergence traced nearly identical trajectories across both the hospital-based Cohorts and the community-based Validation-BC cohort (Extended Figure 2f).

### Longitudinal Consistency and Intra-individual Reproducibility

Next, to evaluate the assay’s clinical applicability and temporal stability, we analyzed the longitudinal Validation-HC cohort (n = 192) (Figure 2a). At baseline, eGFRu formula demonstrated a remarkably strong correlation with eGFRcr (*r* = 0.848) and a negligible mean bias of −1.46 mL/min/1.73m² (Figure 2b, c). The formula exhibited consistently high diagnostic accuracy for identifying all stages of renal impairment, with AUC values uniformly exceeding 0.9 (Figure 2d). This robust performance was maintained in both normoalbuminuric and albuminuric patients (Figure 2e), and exhibited a robust downward trend consistent with increasing CKD severity (Figure 2f). Longitudinal analysis of repeated samples (n = 192 pairs) demonstrated extraordinary temporal consistency (*r* = 0.95), establishing a near-identical “signature” for each individual over observation periods exceeding 1 year (Figure 2g, Supplemental Table 2). Bland-Altman analysis substantiated this, showing a minimal mean bias of -1.27 mL/min/1.73m² with no proportional bias (Figure 2h). In a subset with ≥3 visits (n = 94) (Figure 2i), we found no significant difference in Mean Absolute Error between single measurements and the three-visit average (13.83 vs. 13.59 mL/min/1.73m², P=0.632).

This confirms that a single spot urine sample provides a sufficiently robust estimate. While raw spot urine exhibited substantial intrinsic fluctuations (mean CV: 36.5%), our ratio-based normalization successfully suppressed this hydration-dependent noise (Extended Figure 3a). eGFRu stability was comparable to eGFRcr in low-to-intermediate proteinuria (CV 9.60% vs. 8.12%) but became more variable in the high proteinuria group (≥ 0.5 g/gCr, 17.3%, P < 0.001; Figure 2j). This “Excess CV” correlated positively with proteinuria magnitude and negatively with uCre and eGFRcr (Extended Figure 3b-d). This variance likely arises from a combination of biological factors—such as distal nephron stress induced by heavy glomerular leakage and inherently unstable biomarker excretion in advanced renal decline—and analytical factors, including technical interference from massive proteinuria or mathematical amplification in highly dilute urine.

### Precision Stratification for SGLT2 Inhibitor Therapy via Integrated Clinical Modeling

To evaluate the clinical utility of MUC1_CD9_ for precision medicine, we assessed whether a completely blood-test-free, urine-based algorithm (integrating the MUC1_CD9_ -based eGFRu, ACR, and DM status) could accurately replicate the current blood-based eligibility standards for SGLT2i therapy and high-risk CKD screening.

In patients aged 18–55 years (n=267) (a target demographic for early screening efficacy^34^), the urine-based algorithm demonstrated near-perfect concordance with current blood-based eligibility standards (Accuracy: 96.6%; Kappa: 0.919; PPV: 97.4%; sensitivity: 91.5%; specificity: 98.9%), supporting its potential for blood-test-free prescreening strategy. The scatter plot analysis confirmed that the urine-based eGFR tightly aligned with the standard eGFRcr across the critical clinical decision thresholds (20, 45, and 60 mL/min/1.73 m²), with minimal misclassification (Figure 3a).

**Figure 3.**
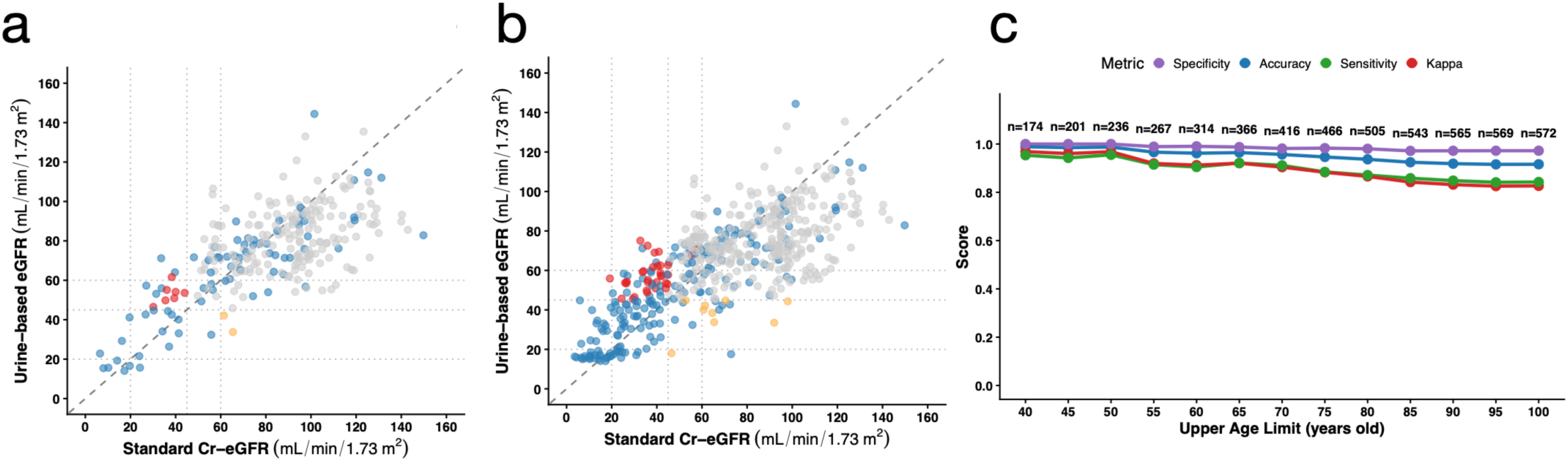
Performance and stability of SGLT2 inhibitor indication criteria based on eGFRu. a, b, Agreement of SGLT2i indication between eGFRu and eGFRcr in young adults (aged 18–55 years) (a) and the full adult cohort (aged 18–85 years) (b); data points are colour-coded by concordance: blue, both positive; grey, both negative; red, eGFRu-positive only; yellow, eGFRcr-positive only. c, Transition of diagnostic accuracy (blue) and Cohen’s κ coefficient (red) as the upper age limit is increased from 40 to 100 years in 5-year increments. SGLT2i, sodium–glucose cotransporter-2 inhibitor; AUC, area under the curve.

Notably, while diagnostic performance gradually declined as the upper age limit was extended to 85 years (n=543), the algorithm maintained robust reliability (Accuracy: 92.4%; Kappa: 0.842; PPV: 95.6%; sensitivity: 85.8%; specificity: 97.2%) (Figure 3b). A trend analysis of these diagnostic metrics revealed that despite a slight decrease in sensitivity among the elderly, overall accuracy and Cohen’s kappa remained exceptionally high across all age subgroups (Figure 3c). Crucially, this consistently high overall performance is driven by a low rate of false positives, suggesting a minimal risk of overprescription. These findings underscore MUC1_CD9_ as a highly direct, stable, parenchyma-derived sensor for clinical prescreening across a broad age spectrum.

### Prognostic Value and Stratification for Precision Therapy

To evaluate the clinical impact of eGFRu on disease progression, we analyzed the longitudinal trajectory of eGFRcr in a prospective cohort (n = 86 in Validation-HC) (Figure 4). This sub-cohort was specifically restricted to patients aged ≥ 40 years with baseline eGFRcr ≥ 15 mL/min/1.73m², as large-scale reference data confirms that physiological eGFRcr decline typically initiates during the fourth decade of life^35^. We defined the eGFR-Gap Ratio as follows:

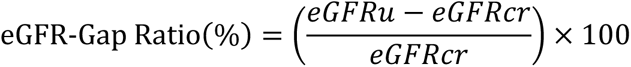

**Figure 4.**
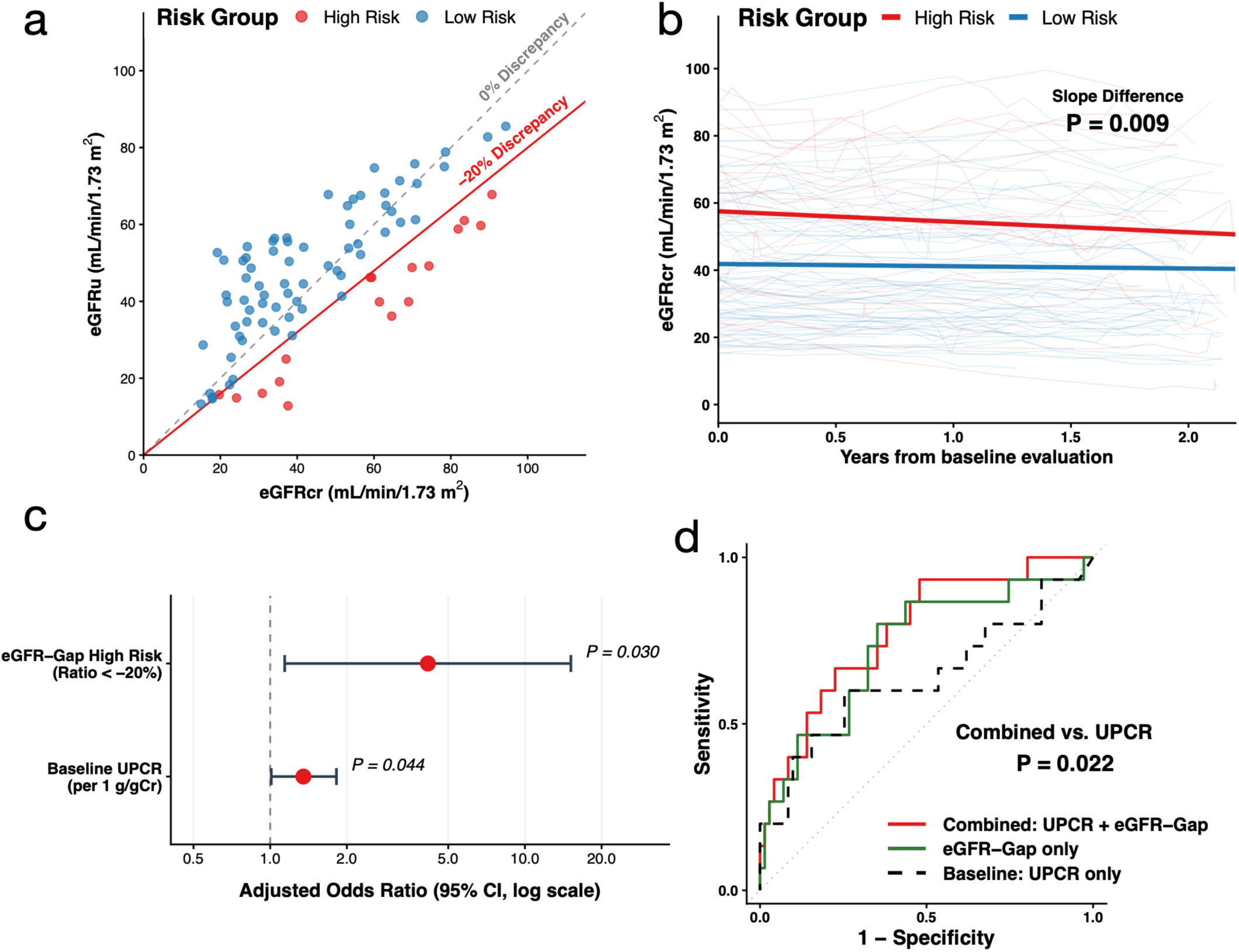
Prognostic utility of the eGFR-Gap Ratio for predicting future kidney function decline. a, Correlation between eGFRu and eGFRcr in the prospective Validation-HC cohort (n = 86); data points are stratified by eGFR-Gap Ratio, with the high-risk group (ratio < −20%) highlighted in red. b, Spaghetti plots of individual longitudinal eGFRcr trajectories stratified by risk group; the high-risk group (red) shows a significantly steeper decline than the low-risk group (blue). c, Forest plot of multivariable logistic regression (Model 2) for predicting rapid renal progression (annualized eGFRcr slope ≤ −5.0 mL/min/1.73 m²/year); adjusted odds ratios (ORs) with 95% confidence intervals are shown for baseline UPCR and eGFR-Gap high-risk status. d, ROC curves evaluating the prognostic performance of the eGFR-Gap Ratio alone (green), baseline UPCR alone (black dashed), and the combined model (red) for identifying rapid progressors (defined as eGFRcr decline > 5 mL/min/1.73 m²/year). P values from linear mixed-effects model (b) and DeLong’s test (d). UPCR, urine protein-to-creatinine ratio.

To compare the longitudinal trajectories of eGFRcr, we employed a linear mixed-effects (LME) model. When stratified by the eGFR-Gap Ratio (the discrepancy between eGFRu and eGFRcr), the High Risk group (Ratio < −20%) (n=17) demonstrated a markedly accelerated decline in kidney function compared to the Low Risk group (mean annual slope: −3.12 vs. −0.67 mL/min/1.73 m²; P interaction = 0.01) (n=69) (Figure 4a, b, Table 2).

**Table 2.**
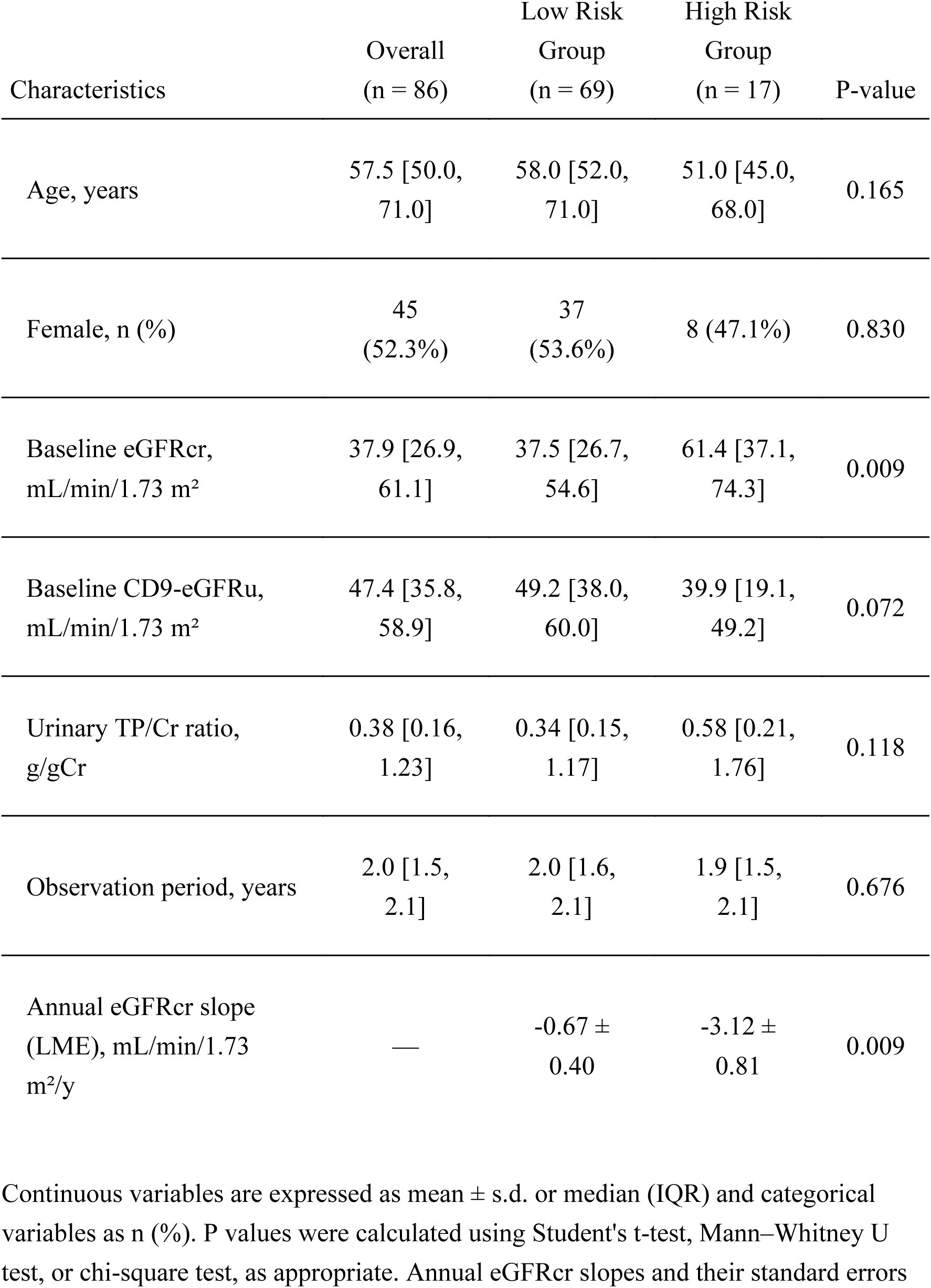
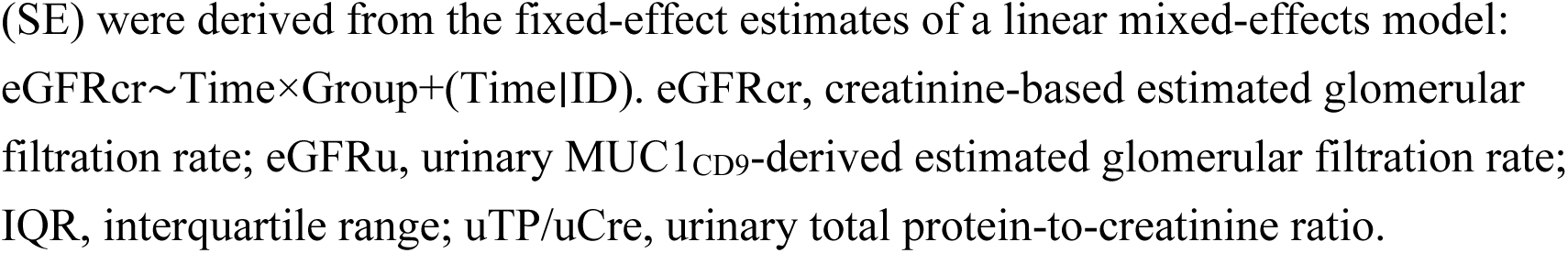
Clinical characteristics of the high-risk and low-risk groups stratified by eGFR-Gap Ratio.

| Characteristics | Overall<br>(n = 86) | Low Risk<br>Group<br>(n = 69) | High Risk<br>Group<br>(n = 17) | P-value |
| --- | --- | --- | --- | --- |
| Age, years | 57.5 [50.0,<br>71.0] | 58.0 [52.0,<br>71.0] | 51.0 [45.0,<br>68.0] | 0.165 |
| Female, n (%) | 45<br>(52.3%) | 37<br>(53.6%) | 8 (47.1%) | 0.830 |
| Baseline eGFRcr,<br>mL/min/1.73 m<sup>2</sup> | 37.9 [26.9,<br>61.1] | 37.5 [26.7,<br>54.6] | 61.4 [37.1,<br>74.3] | 0.009 |
| Baseline CD9-eGFRu,<br>mL/min/1.73 m<sup>2</sup> | 47.4 [35.8,<br>58.9] | 49.2 [38.0,<br>60.0] | 39.9 [19.1,<br>49.2] | 0.072 |
| Urinary TP/Cr ratio,<br>g/gCr | 0.38 [0.16,<br>1.23] | 0.34 [0.15,<br>1.17] | 0.58 [0.21,<br>1.76] | 0.118 |
| Observation period, years | 2.0 [1.5,<br>2.1] | 2.0 [1.6,<br>2.1] | 1.9 [1.5,<br>2.1] | 0.676 |
| Annual eGFRcr slope<br>(LME), mL/min/1.73<br>m<sup>2</sup>/y | — | -0.67 ±<br>0.40 | -3.12 ±<br>0.81 | 0.009 |

Continuous variables are expressed as mean ± s.d. or median (IQR) and categorical variables as n (%). P values were calculated using Student's t-test, Mann–Whitney U test, or chi-square test, as appropriate. Annual eGFRcr slopes and their standard errors
(SE) were derived from the fixed-effect estimates of a linear mixed-effects model: $\text{eGFR}_{\text{cr}} \sim \text{Time} \times \text{Group} + (\text{Time} | \text{ID})$ . eGFR<sub>cr</sub>, creatinine-based estimated glomerular filtration rate; eGFR<sub>u</sub>, urinary MUC1<sub>CD9</sub>-derived estimated glomerular filtration rate; IQR, interquartile range; uTP/uCre, urinary total protein-to-creatinine ratio.

Strikingly, we observed a clinical paradox: High Risk patients presented with significantly higher baseline eGFRcr than Low Risk patients (61.4 vs. 37.5 mL/min/1.73 m², *p* < 0.01) despite their worse future outcomes. This accelerated decline was not attributable to differences in baseline treatment, as the use of renin-angiotensin system and SGLT2 inhibitors was comparable between groups.

To investigate its clinical utility, we defined “rapid progression” as an annualized eGFRcr slope ≤ −5.0 mL/min/1.73 m²/year. In multivariable logistic regression, High Risk status emerged as a strong independent predictor of rapid progression (adjusted OR 4.16, *p* = 0.030), whereas age, sex, and baseline eGFRcr were not significant (Extended Table 4-6, Figure 4c). Furthermore, ROC analysis confirmed that the eGFR-Gap Ratio alone (AUC 0.731) numerically outperformed baseline proteinuria (UPCR, AUC 0.638). Integrating both markers yielded the highest prognostic accuracy (AUC 0.778, *p* = 0.022), highlighting the eGFR-Gap as a critical early warning signal beyond conventional markers (Figure 4d). Importantly, the independent predictive value of the eGFR-Gap High-Risk status was consistently maintained in a sensitivity analysis utilizing the milder progression threshold of ≤ −3.0 mL/min/1.73 m²/year (Extended Table 7).

### Extension of the eGFR Estimation Model to Tim4-Capture platform

Following the initial correlation analysis, we further validated the robustness of the uEV-based eGFR estimation by developing an alternative model using Tim4-captured uEVs. The Tim4-based model—detailed in Supplemental Figure 1 and its legend—yielded a significant correlation with eGFRcr (*r* = 0.72 for all ages; *r* = 0.73 for the under-60 subgroup). Longitudinal prognostic analyses using the Tim4-capture system yielded results consistent with the primary CD9-based platform (Supplemental Figure 2, Supplemental Table 3-6). These findings reinforce the clinical reliability of the eGFRu concept, demonstrating that both CD9- and Tim4-capture systems for MUC1-positive uEVs provide critical prognostic information for identifying patients at risk of rapid renal progression.

### Cellular Basis: snRNA-seq Validates medullary Origin

To elucidate the cellular origin, we integrated snRNA-seq data from the Kidney Precision Medicine Project (KPMP)^36,37^, comparing healthy reference samples (n = 40) with CKD cases (n = 72). The CKD population primarily represented high-priority groups, including diabetic kidney disease (DKD) and hypertension-associated CKD (H-CKD). CD9 and MUC1 were predominantly localized to the collecting duct segments (CCD, OMCD, IMCD) (Extended Figure 4a). To further dissect these changes, we performed a more detailed analysis capable of distinguishing individual cells and patients using the recently expanded human kidney atlas dataset^38^. In CKD, CD9 expression was significantly decreased in specific collecting duct subpopulations, including CCD-PC, dCCD-PC, OMCD-PC, and IC-A. In striking contrast, MUC1 expression was remarkably preserved, showing no evident decrease across any of the collecting duct segments in CKD (Extended Figure 4b).

## Discussion

The findings of this study establish the quantification of CD9/MUC1-positive urinary extracellular vesicles as a prognostic biomarker platform that fundamentally redefines the approach to CKD screening and therapeutic stratification. By leveraging an optimized CD9-capture ELISA to specifically quantify MUC1 on urinary extracellular vesicles, we demonstrated that this assay provides a robust readout of renal parenchymal integrity that surpasses the limitations of conventional blood- and urine-based markers.

### Clinical Robustness and the Mathematical Basis for Stability

A significant hurdle in the clinical implementation of urinary biomarkers is the inherent variability of spot urine concentration. Our mathematical modeling of the eGFRu formula:

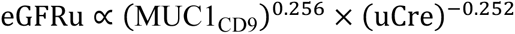

provides a compelling solution to this challenge (Extended Table3). Because both the MUC1-positive vesicles and uCre are subject to the same tubular concentration processes, their fluctuations are synchronized^39^. The near-symmetrical exponents (+0.256 vs. -0.252) effectively neutralize the volume factor, rendering the final estimation virtually independent of the hydration status. Importantly, by functioning effectively as a uCre-normalized ratio, this mathematically derived architecture intrinsically fulfills the latest consensus recommendations of the International Society for Extracellular Vesicles^23,40^, which mandate the normalization of urinary EV metrics to account for highly variable excretion rates.

The robustness of this mathematical architecture is further supported by the finding that the power relationship between eGFRcr and MUC1_CD9_ (∼0.25) is not merely a statistical artifact of overfitting to a specific dataset. When analyzed independently, the optimal exponent for MUC1_CD9_ consistently converged to approximately 0.25 across all three distinct cohorts—the hospital-based Discovery and longitudinal Validation-HC, as well as the community-based Biobank Cohort—maintaining strong explanatory power (*R*² > 0.5) uniformly. This cross-cohort consistency confirms that the derived ∼0.25 exponent reflects a universal biological and mathematical principle of renal parenchymal decline.

This stability is empirically validated by our longitudinal data, which showed an extraordinary intra-individual correlation (*r* = 0.95) and high reproducibility over intervals often exceeding 100 days (Figure 2g, Supplemental Table 2). Unlike raw biomarker measurements that are sensitive to daily metabolic noise, the MUC1_CD9_ to uCre ratio isolates the “intrinsic renal function signal,” enabling reliable long-term monitoring via spot urine without the logistical burden of 24-hour collection.

Furthermore, when directly compared with complex machine learning algorithms (e.g., Random Forest), our simple ratio-based power model demonstrated superior generalizability. While the Random Forest model —trained on independent raw variables (MUC1_CD9_, uCre, and age) without prior ratio calculation—exhibited severe overfitting in the discovery dataset (Pearson’s *r* = 0.954), its performance substantially declined in the independent validation cohort (*r* = 0.837). In contrast, our parsimonious power model maintained robust performance across cohorts, achieving the highest correlation (*r* = 0.849) and diagnostic accuracy for detecting eGFRcr < 60 mL/min/1.73 m² (AUC = 0.935) in the independent validation setting. This underscores that our formula captures the essential biological signal of renal function rather than fitting to noise.

### Resolving the “CKD Blind Spot” and Etiology-Independent Detection

The most immediate clinical contribution of this work is the resolution of the “CKD blind spot“—the failure of standard proteinuria-based screening to identify early-stage renal decline, particularly in non-proteinuric phenotypes like hypertensive nephrosclerosis. Even in patients with eGFRcr already reduced to 30–45 mL/min/1.73m², the absence of significant proteinuria renders these high-risk individuals clinically “invisible,” leading to missed opportunities for timely therapeutic intervention.

Our data confirmed that eGFRu reliably assesses renal impairment even without significant albuminuria/proteinuria. Notably, incorporating urinary total protein (uTP) into the estimation model provided no additional diagnostic benefit, maintaining equivalent or superior AUC values across all stages (Supplemental Figure 3). This highlights that the ratio-based approach captures essential functional information independently of traditional injury markers.

Pathophysiologically, MUC1_CD9_ acts as a proxy for distal nephron integrity: while proteinuria reflects active glomerular injury, the decline in MUC1_CD9_ likely reflects the structural and functional compromise of the collecting duct system. This segment-specific information is particularly transformative when integrated with traditional markers. The synergy between distal nephron integrity (MUC1_CD9_) and glomerular permeability (proteinuria) achieved a near-perfect concordance with standard blood-based criteria for identifying SGLT2i candidates. By unmasking “invisible” yet treatable high-risk patients, this platform opens a decisive therapeutic window for renoprotective intervention when it is most effective.

### Resolving Marker Discordance: Mathematical Resilience to Muscle Mass Bias

The fundamental advantage of our approach lies in capturing the intrinsic discordance between systemic and parenchymal markers.

As demonstrated by the mathematical architecture of standard eGFR equations, large negative exponents render creatinine-based estimates hypersensitive to non-renal factors. Even minor fluctuations in sCre lead to disproportionately large shifts in calculated eGFR, often providing an over-optimistic assessment in individuals with lower muscle mass. By shifting the primary diagnostic signal from a systemic metabolite to a localized parenchymal reporter (MUC1_CD9_), the eGFRu platform bypasses this metabolic “noise”. This mathematical resilience is a key driver of “clinical equity,” ensuring that patients are staged according to their true renal parenchymal integrity rather than their muscular profile—a concept strongly supported by its robust alignment with cystatin C.

### Prognostic Utility of the eGFR-Gap and Longitudinal Observations

As demonstrated by the clinical paradox observed in our longitudinal cohort, standard sCr screening critically fails to identify highly vulnerable patients because their baseline filtration estimates are falsely elevated. The prognostic power of the eGFR-Gap operates independently of both initial filtration-based staging and glomerular protein leakage, serving as a unique indicator of the “parenchymal reserve” available to compensate for ongoing injury. This observation functionally mirrors recent large-scale evidence regarding the discordance between creatinine- and cystatin C-based eGFR (eGFRdiff)^16^. A meta-analysis of over 800,000 individuals demonstrated that a large negative eGFRdiff (where eGFRcys is ≥30% lower than eGFRcr) is a robust predictor of all-cause mortality, cardiovascular events, and kidney failure. By generalizing the concept of “gap analysis” to include urinary vesicle-derived signals, clinicians can proactively identify the high-risk phenotype prone to rapid functional decline.

### Biological Insight: Medullary Integrity and Cellular Plasticity

The medullary collecting duct is often the first site of structural remodeling in response to CKD, transitioning from early qualitative disorganization to late-stage quantitative parenchymal loss. As observed in the single-nucleus RNA-seq (KPMP) analysis^37^, the divergent, bimodal response of MUC1 aligns with recent evidence characterizing it as a critical homeostatic responder to chronic medullary stress^41^ (Importantly, this functional adaptation to stress is conceptually distinct from inherent architectural vulnerabilities driven by MUC1 genetic variants^42,43^.) Because MUC1 transcript levels are maintained at the individual cell level in CKD, the progressive decline in the double-positive uEV pool could involve multiple underlying mechanisms. It may reflect not only the quantitative loss of surviving tubular epithelial cells but also other potential factors. For instance, the down-regulation of CD9 in specific collecting duct segments, as observed in our snRNA-seq analysis, could directly reduce the release of double-positive EVs. Furthermore, stress-induced impairments in apical membrane trafficking and EV secretion dynamics might also play a role. Therefore, the progressive decline in the CD9/MUC1 uEV pool serves as a composite indicator, reflecting both the physical loss of nephrons and their functional exhaustion under chronic stress.

### Scalability and Global Health Impact

A transformative diagnostic tool must be robust under real-world logistics to prevent widening global health inequities^2^. The finding that eGFRu remained stable and quantifiable in samples cryopreserved for over a decade provides critical evidence of the assay’s exceptional analytical stability. This robustness enables retrospective mapping of renal function trajectories using historical biobank specimens, significantly enhancing the biomarker’s utility for large-scale epidemiological research and decentralized clinical monitoring. By lowering the barrier to high-precision monitoring, this platform facilitates a proactive public health approach, potentially reducing global disparities in ESKD incidence by identifying and protecting patients years before they reach the point of no return.

## Limitations

Despite the large multicenter cohort (n = 860), several limitations should be acknowledged. First, the study population was exclusively Japanese; validation in multi-ethnic cohorts is essential to ensure global applicability. Second, while the high consistency with CysC is encouraging, future studies should include “gold standard” comparisons with inulin or 24-hour creatinine clearance. Therefore, the current utility of eGFRu lies primarily in its prognostic precision for identifying future renal decline rather than as a definitive replacement for eGFR measurement. Third, although our platform successfully quantified the MUC1-positive uEV pool, direct physical characterization of these captured vesicles via nanoparticle tracking analysis or electron microscopy remains to be demonstrated in further detail. Finally, while the platform’s scalability is promising, a comprehensive health economic evaluation is needed to determine the cost-effectiveness of integrating this uEV-based assay into targeted population screening programs or decentralized healthcare frameworks.

## Conclusion

The uEV-MUC1 assay expands the capabilities of non-invasive renal diagnostics by shifting the focus from simple filtration estimation to comprehensive risk stratification. By capturing both the molecular signatures of tubular compromise and glomerular protein leakage from a single spot urine sample, this platform provides a completely non-invasive, highly scalable readout that effectively addresses the “CKD blind spot.” Specifically, the eGFR-Gap provides a unique window into the “parenchymal reserve,” allowing for the identification of high-risk individuals prone to rapid functional decline even when their baseline serum creatinine levels appear preserved. This prognostic precision establishes a powerful prescreening strategy in CKD management—moving from the reactive detection of advanced disease to proactive, biology-based prevention and precision intervention at an earlier, more treatable stage.

## Methods

### Study design and participants

This was a multicenter, cross-sectional observational study conducted between April 2022 and December 2025 at the University of Tokyo Hospital (Departments of Pediatrics, Pediatric Surgery, Nephrology and Endocrinology, and Geriatric Medicine) and collaborating pediatric and nephrology centers in Japan, including Hokkaido University Hospital (Department of Pediatrics), Saitama Medical University Hospital (Department of Pediatrics), Chiba Children’s Hospital (Department of Nephrology), Tokyo Women’s Medical University Hospital (Department of Pediatric Nephrology), Toho University Omori Medical Center (Renal Center), Tokyo Metropolitan Bokutoh Hospital (Departments of Pediatrics and Neonatology), Aichi Children’s Health and Medical Center (Department of Nephrology), and Japanese Red Cross Aichi Medical Center Nagoya Daini Hospital (Department of Pediatrics).

The Discovery Cohort (n=471) comprised patients aged ≥16 years from hospital-based clinics, with data collected over a 30-month period between May 25, 2023, and December 5, 2025.

For independent validation, two cohorts were established:

1. The Hospital Cohort (Validation-HC) (n=192): A longitudinal validation set consisting of patients aged ≥16 years whose records were obtained between June 3, 2022, and August 22, 2025 (spanning approximately 39 months).
2. The Biobank Cohort (Validation-BC): To evaluate the general population and provide validation across a broad chronic kidney disease (CKD) spectrum, a community-based validation set (n = 197) was derived from the Tohoku Medical Megabank (TMMB) Project^44^. Samples were specifically extracted from the Tohoku Medical Megabank Organization (ToMMo) Three-Generation Cohort (73K baseline survey, released November 2020)^45^ and the Community-Based Cohort (67K baseline survey, released May 2021)^46^. Specimens for this cohort were collected over a 32-month period between May 21, 2013, and January 5, 2016, based on the following criteria:

Normal Control Group (n = 112): Participants were stratified into three age groups (20–39, 40–59, and 60–79 years). To establish reference values, we analyzed 112 healthy individuals, comprising 58 males and 54 females. The age-specific distribution was: 20–39 years (n=39), 40–59 years (n=36), and 60–79 years (n=37). Inclusion required a baseline eGFRcr of 90–150 mL/min/1.73 m², a urinary albumin-to-creatinine ratio (ACR) < 30 mg/gCr, and negative urinalysis results for protein, occult blood, and glucose. Participants were excluded if they had a history of kidney cancer, chronic nephritis, dialysis, nephrotic syndrome, diabetes (Type 1 or 2), hypertension, systemic lupus erythematosus (SLE), autoimmune/collagen diseases, congenital heart disease, or nephrolithiasis.

Renal Decline Group (n = 85): A total of 85 patients with varying degrees of renal impairment were included 48 males and 37 females). Participants were categorized into nine subgroups based on age and eGFRcr levels (< 30, 30–59, and 60–89 mL/min/1.73 m²). Subgroup sizes ranged from n=6 to 14, with the exception of the 20–39 years/ eGFRcr < 30 group (n=1), consistent with biobank availability.

Urine samples were obtained for routine clinical purposes, and an aliquot of each sample was salvaged for research. After collection, samples were aliquoted and stored at −80 °C in the central laboratory. Samples collected at collaborating sites were either stored at 4 °C and transported to the central laboratory within one week or frozen at −80 °C on site and subsequently transported at 4 °C for permanent storage at −80 °C. Detailed procedures were performed according to the previously published protocol^27,47^. Crucially, all procedures for urine collection, pre-processing, storage, and subsequent EV analysis strictly adhere to the Minimal Information for Studies of Extracellular Vesicles 2023 (MISEV2023) guidelines^21,40^ and the consensus recommendations of the ISEV Urine Task Force^48^.

Clinical information—including height, weight, diagnosis, blood test results (UN, sCre and CysC), and urinalysis (dipstick, sediment, and biochemical data)—was extracted from medical records and entered into an anonymized electronic database.

Renal function was assessed using eGFRcr calculated from sCre. For adults (18 years and older), the JSN eGFRcr formula was used^49^, and for pediatric individuals, the fifth-order Uemura equation was applied^50^. In Validation-BC, we compared it with the Cystatin C-based eGFR (eGFRcysC), calculated using the Japanese standard equations^51^. The study used existing specimens and clinical information; at the University of Tokyo Hospital, an opt-out approach was adopted in lieu of written informed consent, whereas written informed consent was obtained from all participants at other institutions.

The study protocol was approved by the Ethics Committee of the University of Tokyo Hospital (Approval Nos. 11106-(11), 2022330NI, 2021151NI-(3), 2023156NI- (3)). All procedures were performed in accordance with the Declaration of Helsinki.

### Generation of the MUC1 monoclonal antibody (2G4A6)

A novel monoclonal antibody targeting the variable number tandem repeat (VNTR) region of MUC1, corresponding to the epitope recognized by the previously reported VU4H5 antibody, was generated. The production of this monoclonal antibody was commissioned to Cell Engineering Corporation (Osaka, Japan). Briefly, the antigenic peptide (SAPDTRPAPGC) was synthesized (Sigma-Aldrich, St.Louis, MO, USA) and used to immunize B6D2F1/SIc mice twice at a 2-week interval. Spleen lymphocytes were then harvested and fused with myeloma cells to generate hybridomas, which were seeded across four 96-well plates. Culture supernatants were screened for MUC1-specific antibody production using the PS-Capture Exosome ELISA Kit (Anti-Mouse IgG POD, FUJIFILM Wako, Osaka, Japan) according to the manufacturer’s instructions.

Characterization of MUC1 Antibody Reactivity and EV Capture Optimization MUC1 is known for its aberrant expression and glycosylation in various cancers, where its glycans serve as critical biomarkers^52^. The VNTR region, composed of 20 amino acids (GSTAPPAHGVTSAPDTRPAP), is heavily O-glycosylated, and commercial antibodies against this region exhibit varying reactivity depending on the glycan profile ^53^. We evaluated the reactivity of our newly developed clone, 2G4A6, in comparison with the glycosylation-sensitive clone VU4H5 (reactivity attenuated by glycosylation at Thr13) and the glycosylation-independent clone E29^53^.

Furthermore, we compared two uEV capture platforms: the Tim4-based system, which binds phosphatidylserine (PS) present on various EV populations including exosomes, microvesicles, and apoptotic bodies^54^; and the CD9-based system, which targets a classical exosomal marker.

These evaluations were performed using a newly collected pediatric cohort (n = 88), comprising patients with hypoplastic kidneys, asymptomatic hematuria, and normal urinalysis results.

Initial validation on Tim4 plates demonstrated that 2G4A6 reactivity was nearly identical to the glycosylation-independent clone E29 (r = 0.952) and showed high correlation with VU4H5 (r = 0.922), confirming that 2G4A6 recognizes its target sequence independently of VNTR glycosylation. While all antibodies effectively reflected eGFRcr decline, 2G4A6 demonstrated a nearly identical diagnostic trajectory to the established E29 clone, particularly outperforming VU4H5 in detecting advanced renal impairment (AUC > 0.96 vs. 0.88 for eGFRcr < 30). This strong diagnostic and mathematical alignment provides a solid statistical foundation for its use as a robust diagnostic tool.

Further evaluation on CD9-coated plates revealed an extremely high correlation between 2G4A6 and E29 (r = 0.982). Consistent with this biochemical equivalence, both clones demonstrated outstanding diagnostic precision for identifying clinically significant CKD. For the standard diagnostic threshold of eGFRcr < 60 mL/min/1.73 m², the raw urinary values of 2G4A6 and E29 achieved AUCs of 0.937 and 0.948, respectively. Notably, this diagnostic accuracy progressively increased with disease severity, reaching near-perfect AUCs of 0.987 for both clones in detecting advanced renal impairment (eGFRcr < 30 mL/min/1.73 m²). While the discriminatory power was moderate for mild early-stage impairment (eGFRcr < 90 mL/min/1.73 m²; AUC ≈ 0.70), the exceptionally high and nearly identical performance across all severe CKD thresholds underscores 2G4A6 as a highly reliable and diagnostically equivalent alternative to the established E29 clone on the CD9 platform. Consequently, 2G4A6 was selected as the primary detection antibody in combination with CD9-coated plates for all subsequent large-scale analyses.

### Biotin labeling of antibodies

Antibodies were biotinylated using the Biotin Labeling Kit-NH2 (Dojindo, Kumamoto, Japan). Antibodies were mixed with the supplied buffer in centrifugal filter tubes and centrifuged at 8,000 × g for 10 min. The retained antibodies were reacted with NH2-reactive biotin dissolved in DMSO at 37 °C for 10 min. After three washes with buffer, the purified biotin-labeled antibodies were mixed with buffer and 50% glycerol and stored at −20 °C until use.

### ELISA using Tim4- and CD9-coated plates

Quantification of uEV-associated markers was performed using two commercially available sandwich ELISA platforms: the PS-Capture Exosome ELISA Kit (Streptavidin; Tim4-coated plate, hereafter Tim4 plate) and the CD9-Capture Exosome ELISA Kit (Streptavidin; anti-CD9 antibody–coated plate, hereafter CD9 plate) (both from FUJIFILM Wako). Detailed procedures were performed according to the previously published protocol^47^.

Thawed urine samples were centrifuged at 1,200 × g for 5 min to remove cellular debris and precipitated salts. After three washes with the kit wash buffer, diluted urine samples or dilution buffer (blank) were added to the wells and incubated on a microplate shaker at 550 rpm for 2 h at room temperature. Plates were then washed three times, and biotin-labeled detection antibodies were added and incubated for 1 h with shaking. After five washes, HRP-conjugated streptavidin was added and incubated for 1 h. Following five additional washes, TMB substrate was added for 30 min and the reaction was stopped with stop solution. Absorbance was measured immediately at 450 nm and 620 nm. Final values were calculated as (A450 − A620) minus the corresponding blank value. For cohorts measured within a single assay run, raw absorbance values were used; for cohorts requiring multiple assay days, values were normalized to a pooled healthy adult urine standard included on each plate. To definitively standardize MUC1 quantification across all assays and cohorts, we established a unified Reference Unit (RU). Specifically, the mean MUC1_CD9_ level of healthy young adults (aged < 40 years with eGFRcr ≥ 90 mL/min/1.73 m²) in the Hospital Cohort was defined as 100 RU. All quantified MUC1 values were subsequently converted into and expressed as these Reference Units.

Four ELISA configurations were evaluated on Tim4- and CD9-coated plates: Tim4-capture with CD9 or MUC1 detection, and CD9-capture with CD9 or MUC1 detection. For the main analyses using uEV-based assays, MUC1_Tim4_ (Tim4-capture/MUC1-detection) and MUC1_CD9_ (CD9-capture/MUC1-detection) were selected because they showed the best correlations with eGFRcr.

### Standardization of urinary EV quantification

Because urinary measurements are strongly influenced by urine dilution, uEV markers were analyzed both as raw values and after normalization to urinary creatinine (uCre). uCre was measured in routine clinical laboratories. Based on previous reports, samples with extreme dilution or concentration (uCre < 22.6 mg/dL, or < 10 mg/dL in children, or > 300 mg/dL) were excluded from the statistical analyses^55,56^. For each ELISA configuration, we defined variables such as MUC1_CD9_ (raw) and MUC1_CD9_/uCre (uCre-normalized) and evaluated both formats in correlation and regression models.

### Measurement of Urinary Biomarkers

Concentrations of urinary free MUCIN 1 (ufMUC1) and urinary neutrophil gelatinase-associated lipocalin (uNGAL) were measured using commercially available enzyme-linked immunosorbent assay (ELISA) kits. Specifically, ufMUC1 was quantified using the Human MUCIN 1/MUC1 ELISA Kit (Catalog Number: EHMUC1; Invitrogen, Thermo Fisher Scientific, Waltham, MA, USA), and uNGAL was quantified using the Human NGAL ELISA Kit (Catalog Number: KE00327; Proteintech Group, Rosemont, IL, USA). All assays were performed according to the manufacturers’ instructions.

Absorbance was measured at 450 nm using a microplate reader, and the concentrations were calculated based on standard curves.

### Single-nucleus RNA-seq analysis

Single-nucleus RNA-seq (snRNA-seq) data from the Kidney Precision Medicine Project were used to characterize CD9 and MUC1 expression in human kidneys. Publicly available datasets comprising healthy and CKD specimens were downloaded^37^, processed using standard pipelines (quality control, normalization, clustering, and cell-type annotation), and interrogated for AQP2, CD9 and MUC1 expression across nephron segments. Dot plots and aggregate expression analyses were used to localize expression to distal nephron and collecting duct segments and to compare expression patterns between healthy and CKD kidneys. For detailed quantitative comparisons between healthy reference and CKD specimens, processed datasets were downloaded directly from the platform^38^. A representative subset of 20,000 cells was extracted and analyzed using R software. The processed data—which included quality control, normalization, clustering, and cell-type annotation performed by the consortium—were interrogated to generate box plots comparing CD9 and MUC1 expression levels across distinct functional and disease states. To account for inter-individual variability and avoid pseudoreplication, a pseudo-bulk approach was applied; single-cell expression data were aggregated by calculating the mean expression level per cell type for each individual donor. These donor-based mean expression values were then utilized to generate box plots and perform statistical comparisons between healthy and CKD groups using the Wilcoxon rank-sum test.

### Statistical analysis

#### 1. Software and Environment

All statistical analyses and data visualizations were performed using R version 4.5.2 (R Foundation for Statistical Computing, Vienna, Austria). Data manipulation and plotting were primarily conducted using the tidyverse suite (including dplyr, tidyr, and ggplot2). Linear mixed-effects (LME) models were implemented using the lme4 and lmerTest packages, with P-values for fixed effects estimated via Satterthwaite’s degrees of freedom approximation. Diagnostic performance and receiver operating characteristic (ROC) analysis were performed using the pROC package.

#### 2. Statistical Modeling of Urinary Biomarker Determinants

To identify independent clinical correlates of MUC1_CD9_, multivariable linear regression models were constructed to estimate the independent contribution of each predictor while adjusting for confounding factors such as eGFRcr, uCre age, sex, height and weight. To directly compare the relative impact of predictors measured in different units, all continuous variables were standardized (Z-score transformed) prior to analysis, allowing for the calculation of standardized regression coefficients (β). These β values were used to identify the dominant clinical determinants for each biomarker.

Multicollinearity was assessed using the variance inflation factor (VIF); all variables had VIF values < 3.0.

#### 3. Establishment of the MUC1-based eGFR (eGFRu) Formula

The MUC1-based estimated glomerular filtration rate (eGFRu) was established through multi-variable regression modeling using data from the discovery cohort. To handle samples with MUC1 levels below the detection limit or recorded as zero, a minimal offset (representing half of the observed minimum positive value for each platform) was applied prior to logarithmic transformation. Specifically, an offset of 0.1127 was used for MUC1_CD9_, and an offset of 0.0570 was applied for MUC1_Tim4_.

#### 4. Diagnostic Performance and ROC Analysis

The diagnostic accuracy of eGFRu was evaluated using Receiver Operating Characteristic (ROC) curves and Area Under the Curve (AUC) calculations. The performance was validated against standard creatinine-based eGFR (eGFRcr) and cystatin C-based eGFR (eGFRcysC) at clinically relevant thresholds of < 30, < 45, < 60, and < 90 mL/min/1.73 m². The optimal cutoff points were identified using Youden’s Index. Statistical comparisons between different AUC values were performed using DeLong’s test.

#### 5. Longitudinal Analysis using Linear Mixed-Effects (LME) Models eGFR-Gap Ratio and Selection of Clinical Threshold

To evaluate the predictive capacity of eGFRu for future renal function decline, we conducted a prospective longitudinal analysis in a subset of the Hospital Cohort (n = 86). The primary prognostic indicator was the eGFR-Gap Ratio, defined as the percentage discrepancy between the uEV-derived eGFR and the serum creatinine-based eGFR:

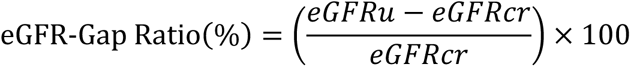

The optimal clinical threshold for the eGFR-Gap Ratio was determined through a sensitivity analysis evaluating cutoffs ranging from −5 to −25. While statistical optimization (maximal Youden index) suggested a threshold of −7.6, we prioritized clinical robustness and the clear differentiation of longitudinal slopes. The threshold of -20 (%) for the eGFR-Gap Ratio was selected based on sensitivity analysis to identify a high-risk subgroup (representing approximately 20% of the cohort) where the divergence in future annualized eGFRcr slopes was most statistically significant.

This threshold was prioritized for its clinical utility in identifying the specific phenotype most prone to rapid functional deterioration.

### Longitudinal Trajectory and Outcome Modeling

To compare the longitudinal trajectories of eGFRcr between the High-Gap and Low-Gap risk groups, we employed a linear mixed-effects (LME) model. This model accounted for repeated measures and intra-individual correlation by incorporating individual random intercepts and random slopes for each participant. The model included fixed effects for time (years from baseline), risk group (High-Gap Risk vs. Low-Gap Risk), and their interaction term (Time×Group). The reported annual rates of eGFRcr decline (slopes) and their corresponding standard errors (SE) were derived from the fixed-effect estimates of the interaction model. Multivariable LME models were further adjusted for baseline proteinuria (Urinary TP/Cr ratio) to confirm the independent predictive value of the eGFR-Gap Ratio.

### Prediction of Rapid Renal Decline

“Rapid renal decline” was defined as an annualized eGFRcr slope of ≤−5.0 mL/min/1.73 m²/year (or the lowest 15th percentile). To identify factors associated with rapid progression, we performed multivariable logistic regression analysis, adjusting for age, sex, baseline eGFRcr, and proteinuria (UPCR). As a sensitivity analysis, we also evaluated an alternative threshold for moderate disease progression, defined as an annualized eGFRcr slope of ≤ −3.0 mL/min/1.73 m²/year. Finally, diagnostic performance for predicting rapid decline was assessed via ROC analysis, comparing the eGFR-Gap Ratio against baseline UPCR.

### SGLT2i Eligibility and Clinical Utility Analysis

The reference standard for SGLT2i therapy and high-risk CKD screening was defined in accordance with the 2024 KDIGO CKD guidelines:

- Type 2 diabetes with CKD: eGFRcr < 60 mL/min/1.73 m² or ACR ≥ 30 mg/g.
- Non-diabetic CKD: eGFRcr ≤ 45 mL/min/1.73 m² (irrespective of ACR) or ACR ≥ 200 mg/g (for those with eGFRcr > 45 mL/min/1.73 m²).

Importantly, patients with an eGFRcr < 20 mL/min/1.73 m² were deliberately retained in the analysis. Although de novo SGLT2i initiation may not be strictly recommended at this advanced stage, detecting such severe renal impairment remains clinically imperative for comprehensive screening and timely nephrology referral. Heart failure–based indications were not incorporated because heart failure status was not systematically captured in this cohort.

The performance of the urine-based algorithm was evaluated using confusion matrices to calculate overall accuracy, Cohen’s Kappa coefficient, sensitivity, specificity, and positive predictive value (PPV). Furthermore, an age-stratified sensitivity analysis was conducted by progressively increasing the upper age limit of the cohort (from ≤55 up to ≤85 years) to evaluate the robustness of the algorithm against age-related metabolic variations.

### Selection of the Clinical Threshold

The optimal clinical threshold for the eGFR-Gap Ratio was determined through a sensitivity analysis evaluating cutoffs ranging from −5 to −25. While statistical optimization (maximal Youden index) suggested a threshold of −7.6, we prioritized clinical robustness and the clear differentiation of longitudinal slopes. The threshold of - 20 (%) for the eGFR-Gap Ratio was selected based on sensitivity analysis to identify a high-risk subgroup (representing approximately 20% of the cohort) where the divergence in future annualized eGFRcr slopes was most statistically significant. This threshold was prioritized for its clinical utility in identifying the specific phenotype most prone to rapid functional deterioration.

## Acknowledgement

We express our deepest gratitude to all the patients and participants who made this study possible. We also thank Haru Kato, Aiko Kitamura, and Yoshie Kikuchi (The University of Tokyo) for their technical support.

## Author contributions

Yutaka.H. designed the study, analyzed data, and wrote the manuscript. T.N. performed experiments, contributed to drafting the manuscript, and collected clinical samples and patient data. Yosuke.H., K.T, J F, S.O., Y.K., S.K., R.K., Tae.O., Yuko.H, Y.G., K.M., N.F., Takayuki. O., M.H., collected clinical samples and patient data. M.N, and M.K. supervised the work, and revised the manuscript.

All authors read and approved the final version of the manuscript.

## Data Availability

Individual patient clinical datasets analyzed during the current study are not publicly available and cannot be shared outside the participating institutions. This restriction is strictly enforced by the opt-out consent protocol approved by the Ethics Committees, which mandates that all de-identified data must be securely stored within the institution to prevent external leakage and protect patient privacy. Aggregate or summary data that do not identify individuals are included in this published article and its supplementary information files, or may be available from the corresponding author upon reasonable request.

---

**Extended Figure 1.**
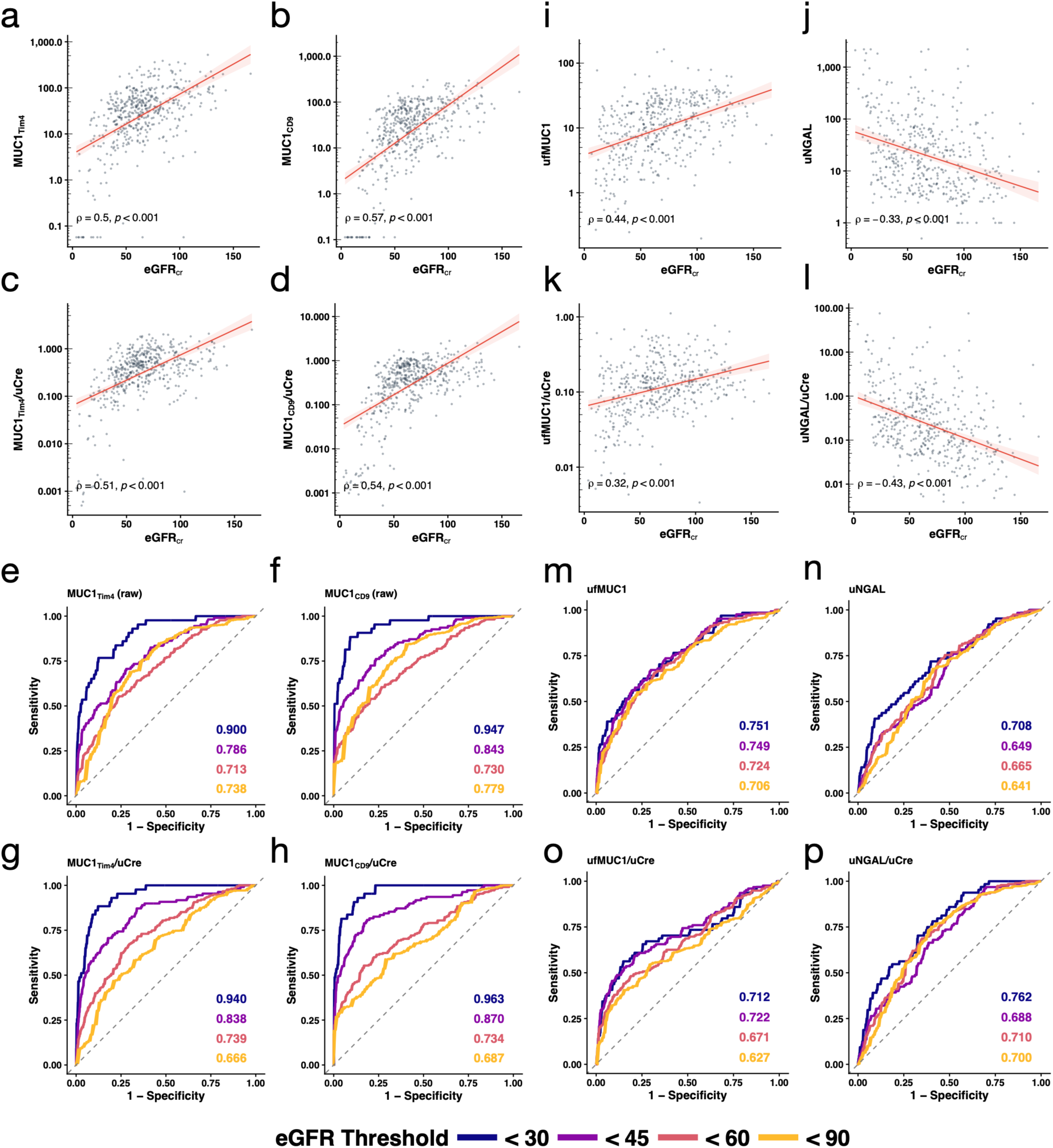
Correlation Analysis and Diagnostic Performance of Raw and Creatinine-Normalized Urinary MUC1-related Markers for Identifying Impaired Renal Function. a-d, Scatter plots showing the correlation between eGFRcr and (a) raw urinary MUC1(Tim4)(RU), (b) raw urinary MUC1(CD9)(RU), (c) creatinine-normalized urinary MUC1(Tim4)(RU), and (d) creatinine-normalized urinary MUC1(CD9)(RU) in the Discovery cohort. The Y-axis is presented on a log10 scale. The red lines represent linear regression fits with 95% confidence intervals (shaded areas). Statistical significance was assessed using Spearman’s rank correlation coefficient (ρ). eGFRcr, creatinine-based estimated glomerular filtration rate; uCre, urinary creatinine. e-h, Receiver operating characteristic (ROC) curves of (e) raw MUC1(Tim4), (f) raw MUC1(CD9), (g) creatinine-normalized MUC1(Tim4), and (h) creatinine-normalized MUC1(CD9) for identifying different eGFRcr thresholds (< 30, 45, 60, and 90 mL/min/1.73 m2). AUC values for each threshold are indicated within the panels. The diagonal dashed line represents the line of identity. Color-coded curves represent the respective eGFRcr cut-offs. AUC, area under the curve. i-l, Scatter plots showing the correlation between eGFRcr and conventional soluble urinary biomarkers: (i) raw free urinary MUC1 (ufMUC1), (j) raw urinary NGAL (uNGAL), (k) creatinine-normalized ufMUC1, and (l) creatinine-normalized uNGAL. The Y-axis is presented on a log10 scale. The red lines represent linear regression fits with 95% confidence intervals (shaded areas). Statistical significance was assessed using Spearman’s rank correlation coefficient (ρ). m-p, Receiver operating characteristic (ROC) curves of (m) raw ufMUC1, (n) raw uNGAL, (o) creatinine-normalized ufMUC1, and (p) creatinine-normalized uNGAL for identifying different eGFRcr thresholds. AUC values for each threshold are indicated within the panels. The diagonal dashed line represents the line of identity. Color-coded curves represent the respective eGFRcr cut-offs. ufMUC1, free urinary MUC1; uNGAL, urinary neutrophil gelatinase-associated lipocalin.

**Extended Figure 2.**
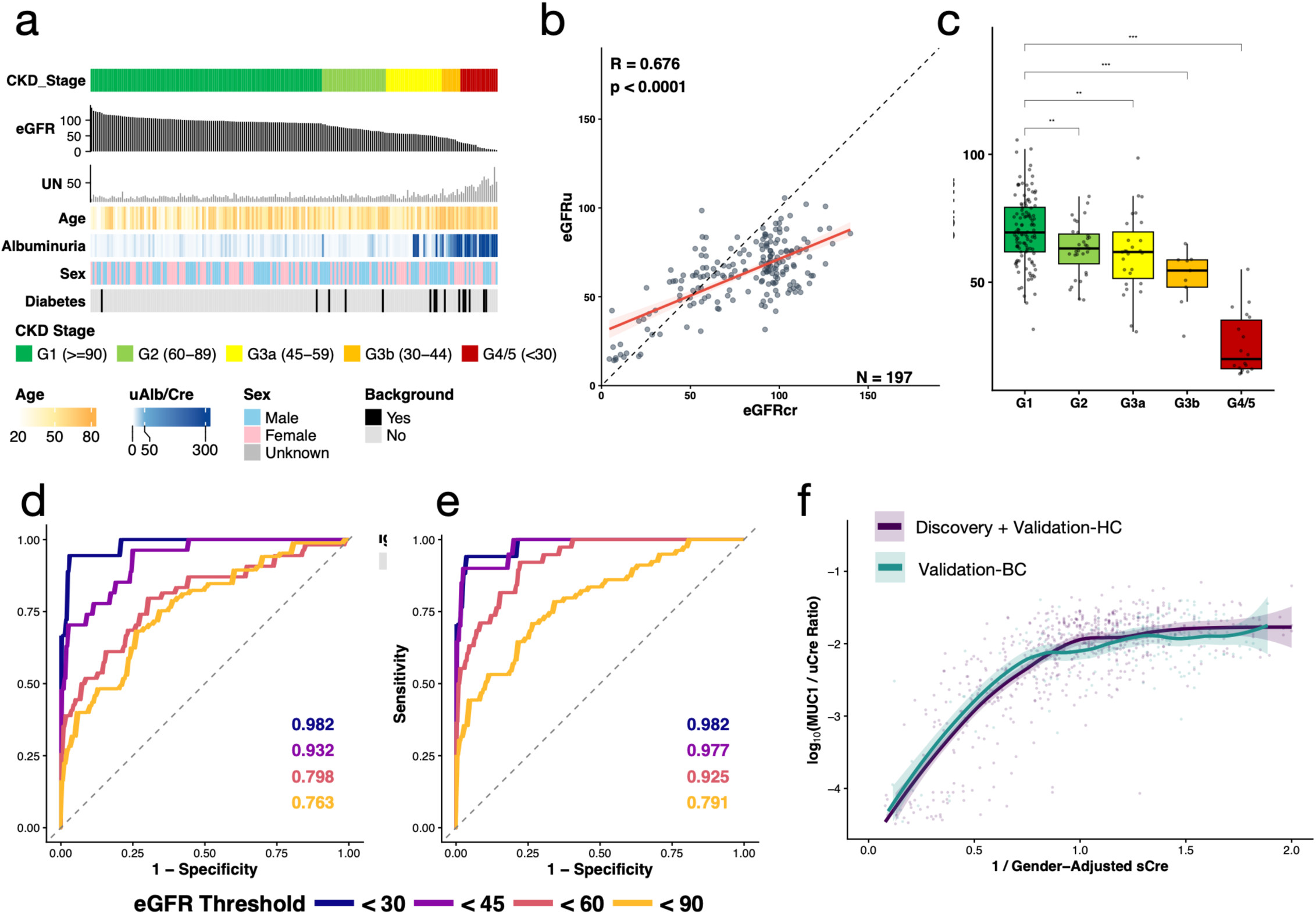
External validation and biological characterization of eGFRu in the biobank cohort. a, Clinical and molecular landscape of the external biobank cohort (Validation-BC; n = 197). b, Correlation between eGFRu and eGFRcr in the biobank cohort. The correlation between eGFRcr and eGFRu was assessed using Pearson’s correlation coefficient. c, The box-and-whisker plots represent the distribution of eGFRu, categorized by clinical CKD stages (G1 to G4/5) based on reference eGFRcr. Statistical significance was determined using the Wilcoxon rank-sum test, comparing each stage to the G1 group (\**p* < 0.01, \*\*\**p* < 0.001). d, e, ROC curves of eGFRu for identifying stages of renal impairment defined by eGFRcr (d) and eGFRcysC (e) thresholds (< 30, 45, 60, and 90 mL/min/1.73 m²). f, Scatter plot of log-transformed MUC1/uCre against the inverse of sex-adjusted urinary creatinine (1/sex-adjusted uCre) for all subjects aged ≥18 years; data from the discovery and Validation-HC cohorts (purple) and the Validation-BC cohort (green) are shown with LOESS regression lines and shaded areas. ROC, receiver operating characteristic.

**Extended Figure 3.**
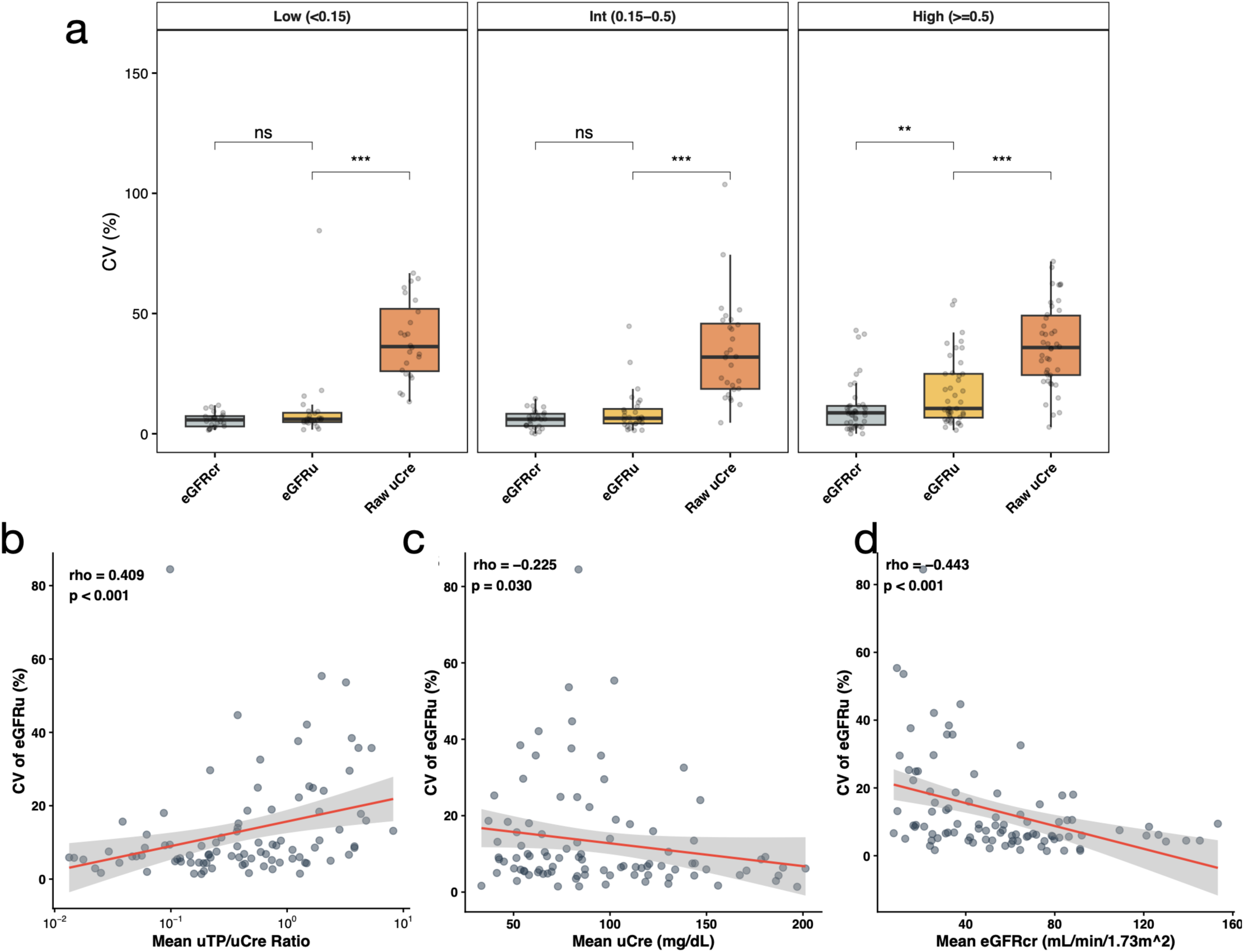
Factors associated with the intra-individual stability of eGFRu. a, Intra-individual coefficients of variation (CV) of eGFRcr (grey) and eGFRu (orange) in patients with ≥3 independent measurements (n = 102), stratified by mean urinary total protein-to-creatinine ratio (uTP/uCre): Low (< 0.15 g/gCr), Intermediate (0.15–0.5 g/gCr), and High (≥ 0.5 g/gCr). Boxes represent the interquartile range (IQR) with the median; whiskers extend to 1.5× IQR; individual data points are superimposed. \*\*\**p* < 0.001. b–d, Correlation between CV of longitudinal eGFRu measurements and mean uTP/uCre (b) mean urinary creatinine (uCre; proxy for muscle mass and urine concentration), log-transformed (c), and mean eGFRcr (d), in patients with ≥ 3 visits (n = 102). Scatter plots include linear regression lines with 95% confidence intervals (shaded areas). *P* values from paired Wilcoxon signed-rank test (a) and Spearman’s rank correlation (b–d).

**Extended Figure 4.**
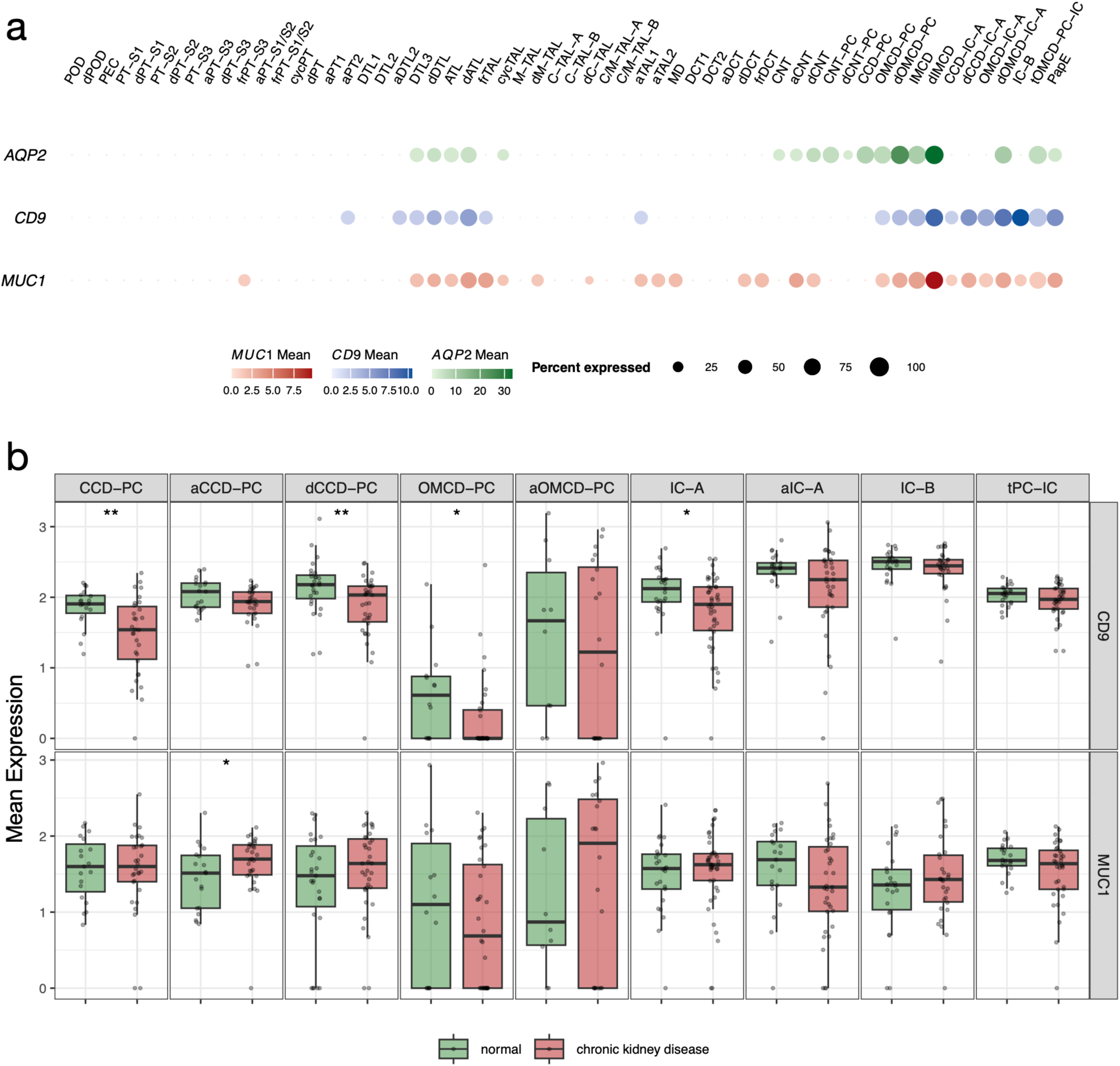
Single-nucleus RNA-seq analysis of CD9 and MUC1 expression in human kidney segments. a, Dot plot of AQP2 (green), MUC1 (blue), and CD9 (red) expression across nephron segments from single-nucleus RNA-sequencing (snRNA-seq) data of the Kidney Precision Medicine Project (KPMP). b, Differential expression of CD9 and MUC1 between healthy reference and chronic kidney disease (CKD) samples in collecting duct cells. Box plots indicate the median and interquartile range of pseudo-bulk mean expression levels, with individual patient (donor) samples overlaid as jittered points. Colors represent the disease status: green (healthy reference) and red (CKD). Statistical significance was determined by the Wilcoxon rank-sum test using donor-level aggregated data; *p<0.05, **p<0.01 (relative to healthy reference).

**Extended Table 1.** Baseline characteristics of the discovery and validation cohorts. Values are expressed as mean ± s.d., median (IQR), or n (%). eGFRcr, creatinine-based estimated glomerular filtration rate; eGFRu, urinary MUC1-derived estimated glomerular filtration rate; HC, hospital-based validation cohort; IQR, interquartile range; NA, not available; uTP/uCre, urinary total protein-to-creatinine ratio.

| Parameter | Discovery (HC) | Validation (HC) | Validation (BC) |
| --- | --- | --- | --- |
| n | 471 | 192 | 197 |
| Age (years) | 57.8 ± 26.8 | 45.8 ± 22.9 | 51.0 ± 14.9 |
| Male, n (%) | 202 (42.9%) | 99 (51.6%) | 106 (53.8%) |
| eGFRcr (mL/min/1.73 m<sup>2</sup>) | 62.3<br>[45.7, 80.0] | 58.8<br>[30.7, 86.1] | 92.0<br>[58, 100] |
| eGFRu (mL/min/1.73 m<sup>2</sup>) | 62.2<br>[49.2, 74.1] | 56.0<br>[39.2, 83.1] | 64.0<br>[53.6, 74.5] |
| Urinary Creatinine (mg/dL) | 83.1<br>[49, 134.5] | 83.6<br>[52.75, 146.3] | 77.0<br>[50, 122] |
| Urinary TP/Cr Ratio (g/gCr) | 0.11<br>[0.04, 0.41] | 0.33<br>[0.09, 1.29] | NA |

**Extended Table 2.**
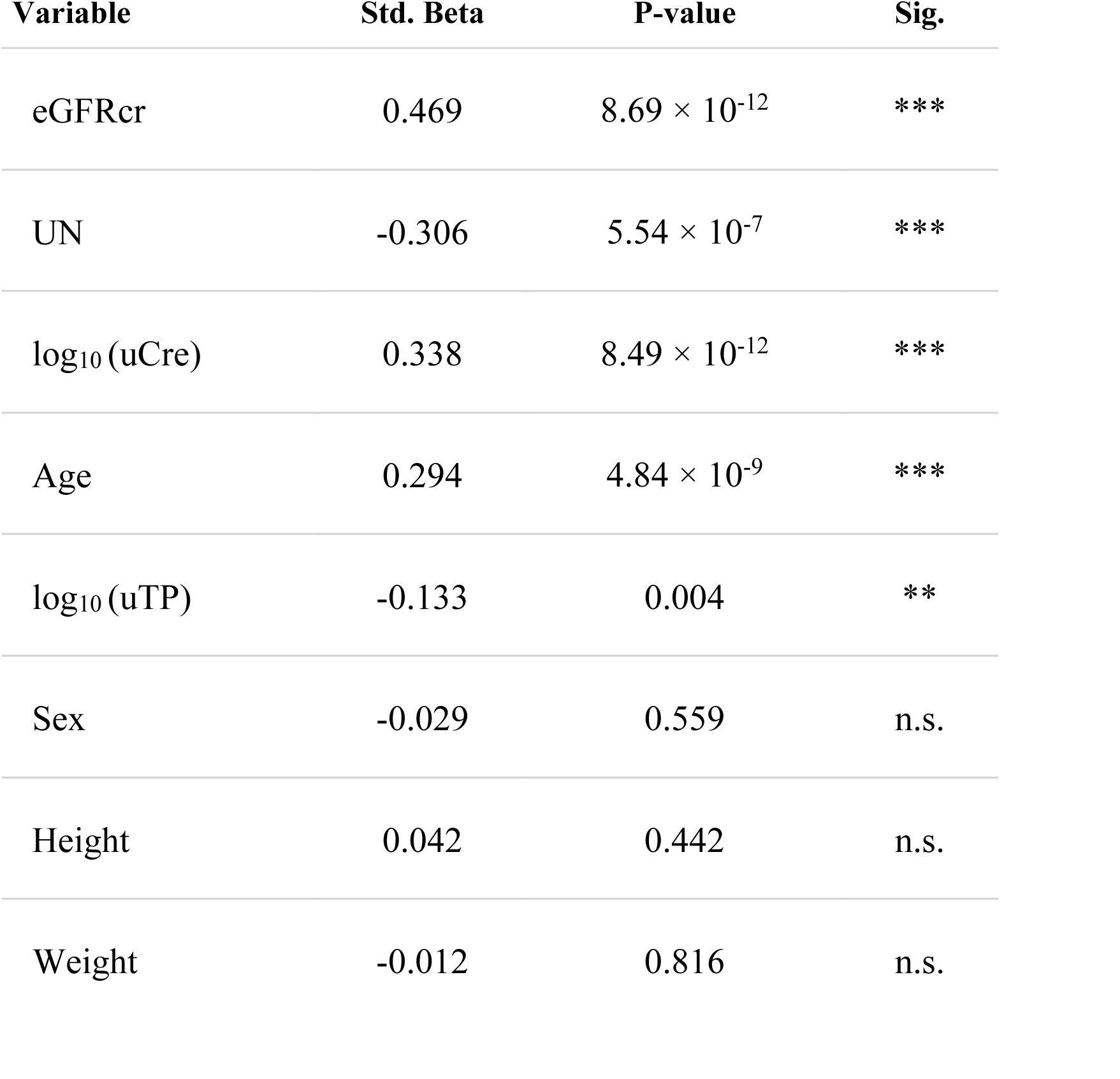
Multivariable regression model for urinary MUC1_CD9_ levels. The dependent variable was log₁₀-transformed MUC1_CD9_; (log₁₀ MUC1_CD9_; n = 163, representing subjects with complete data for all included variables). Independent variables included age, sex, height, weight, eGFRcr, urea nitrogen, log₁₀-transformed urinary creatinine, and log₁₀-transformed urinary total protein. Standardized β coefficients indicate the relative contribution of each predictor. Adjusted *R²* = 0.7508. \*\*\**p* < 0.001; ** *p* < 0.01; * *p* < 0.05; n.s., not significant. eGFRcr, creatinine-based estimated glomerular filtration rate; log-uCre, log₁₀-transformed urinary creatinine; log-uTP, log₁₀-transformed urinary total protein; UN, urea nitrogen.

| Variable | Std. Beta | P-value | Sig. |
| --- | --- | --- | --- |
| eGFRcr | 0.469 | $8.69 \times 10^{-12}$ | *** |
| UN | -0.306 | $5.54 \times 10^{-7}$ | *** |
| log<sub>10</sub>(uCre) | 0.338 | $8.49 \times 10^{-12}$ | *** |
| Age | 0.294 | $4.84 \times 10^{-9}$ | *** |
| log<sub>10</sub>(uTP) | -0.133 | 0.004 | ** |
| Sex | -0.029 | 0.559 | n.s. |
| Height | 0.042 | 0.442 | n.s. |
| Weight | -0.012 | 0.816 | n.s. |

**Extended Table 3.** Multivariable linear regression model for eGFRcr estimation. The model includes natural log-transformed age, urinary creatinine, and urinary MUC1_CD9_ as predictors (n = 471). Sex was coded as male = 1 and female = 0. Adjusted R² = 0.641. eGFRu, urinary MUC1-derived estimated glomerular filtration rate; ln, natural logarithm; MUC1_CD9_, urinary extracellular vesicle MUC1 marker; uCre, urinary creatinine.

| Variable | Coefficient (B) | Std. Error | t value | P value |
| --- | --- | --- | --- | --- |
| Intercept | 2.3656 | 0.0740 | 31.9868 | $2.64 \times 10^{-134}$ |
| $\log_{10}$ (Age) | -0.2763 | 0.0252 | -10.9479 | $5.59 \times 10^{-25}$ |
| $\log_{10}$ (uCre) | -0.2518 | 0.0283 | -8.8843 | $1.41 \times 10^{-17}$ |
| $\log_{10}$ (MUC1<sub>CD9</sub>) | 0.2559 | 0.0099 | 25.7233 | $1.80 \times 10^{-91}$ |
| Sex (Male) | -0.0003 | 0.0139 | -0.0190 | 0.9848 |

**Extended Table 4.** Multivariable logistic regression for predictors of rapid renal progression using eGFR-Gap Ratio as a continuous variable. Rapid progression was defined as an annualized eGFRcr slope of ≤ −5.0 mL/min/1.73 m²/year (n = 15); individual slopes were estimated using a linear mixed-effects model from the eGFR-Gap assessment date. Adjusted odds ratios (ORs) with 95% confidence intervals (CIs) are shown for a model adjusted for age, sex, baseline eGFRcr, and baseline UPCR. CI, confidence interval; eGFRcr, creatinine-based estimated glomerular filtration rate; UPCR, urinary protein-to-creatinine ratio.

| Predictor | Adjusted Odds Ratio (95% CI) | *P*-value |
| --- | --- | --- |
| eGFR-Gap Ratio | 0.108 (0.010–0.755) | 0.040 |
| Age | 1.000 (0.945–1.057) | 0.986 |
| Sex (Female) | 1.938 (0.498–8.502) | 0.351 |
| Baseline eGFRcr | 1.004 (0.970–1.042) | 0.822 |
| Baseline UPCR | 1.525 (1.105–2.249) | 0.020 |

**Extended Table 5.** Baseline clinical characteristics of rapid and non-rapid progressors. Rapid progression was defined as an annualized eGFRcr slope of ≤ −5.0 mL/min/1.73 m²/year from the date of urinary MUC1_CD9_ measurement. Continuous variables are expressed as mean ± s.d. or median (IQR) and categorical variables as n (%). P values were calculated using Student’s t-test, Mann–Whitney U test, or chi-square test, as appropriate. eGFRcr, creatinine-based estimated glomerular filtration rate; IQR, interquartile range; UPCR, urinary protein-to-creatinine ratio.

| Variables | Non-Rapid Progressor<br>(n = 71) | Rapid Progressor<br>(n = 15) | P-value |
| --- | --- | --- | --- |
| **Demographics** |  |  |  |
| Age, years | 59 [51.5, 70] | 52 [45.0, 69.5] | 0.248 |
| Male, n (%) | 35 (49.3%) | 6 (40.0%) | 0.711 |
| **Renal Function** |  |  |  |
| Baseline eGFR<sub>cr</sub>,<br>mL/min/1.73 m<sup>2</sup> | 37.9 [26.7, 59.5] | 48.1 [30.9, 71.4] | 0.292 |
| Baseline UPCR, g/gCr | 0.3 [0.2, 1] | 1.1 [0.2, 2.3] | 0.095 |
| **eGFR-Gap Biomarker** |  |  |  |
| eGFR-Gap Ratio | 0.085 [-0.105, 0.366] | -0.093 [-0.353, -0.049] | 0.005 |
| High Risk<br>(Gap Ratio < -20%), n (%) | 10 (14.1%) | 7 (46.7%) | 0.009 |

**Extended Table 6.** Multivariable logistic regression models for predictors of rapid renal progression using High-Risk categorization. Rapid progression was defined as an annualized eGFRcr slope of ≤ −5.0 mL/min/1.73 m²/year from the date of urinary MUC1_CD9_ measurement. The high-risk group was defined as an eGFR-Gap Ratio of < −20%. Baseline UPCR and eGFRcr were entered as continuous variables. Adjusted ORs with 95% CIs are shown. CI, confidence interval; eGFRcr, creatinine-based estimated glomerular filtration rate; OR, odds ratio; UPCR, urinary protein-to-creatinine ratio.

| **Variable** | **Model 1: OR**<br>**(95% CI)** | **Model 1: P-value** | **Model 2: OR**<br>**(95% CI)** | **Model 2: P-value** |
| --- | --- | --- | --- | --- |
| **High Risk (Gap < -20%)** | **3.99 (0.94-17.19)** | **0.058** | **4.16 (1.11-15.28)** | **0.030** |
| Age, per year | 1.00 (0.95- 1.06) | 0.993 | - | - |
| Sex (Female) | 2.01 (0.51- 9.03) | 0.329 | - | - |
| Baseline eGFRcr, per unit | 1.01 (0.97- 1.05) | 0.677 | - | - |
| **Baseline UPCr, per g/gCr** | **1.43 (1.03- 2.06)** | **0.033** | **1.35 (1.03- 1.88)** | **0.044** |

**Extended Table 7.**
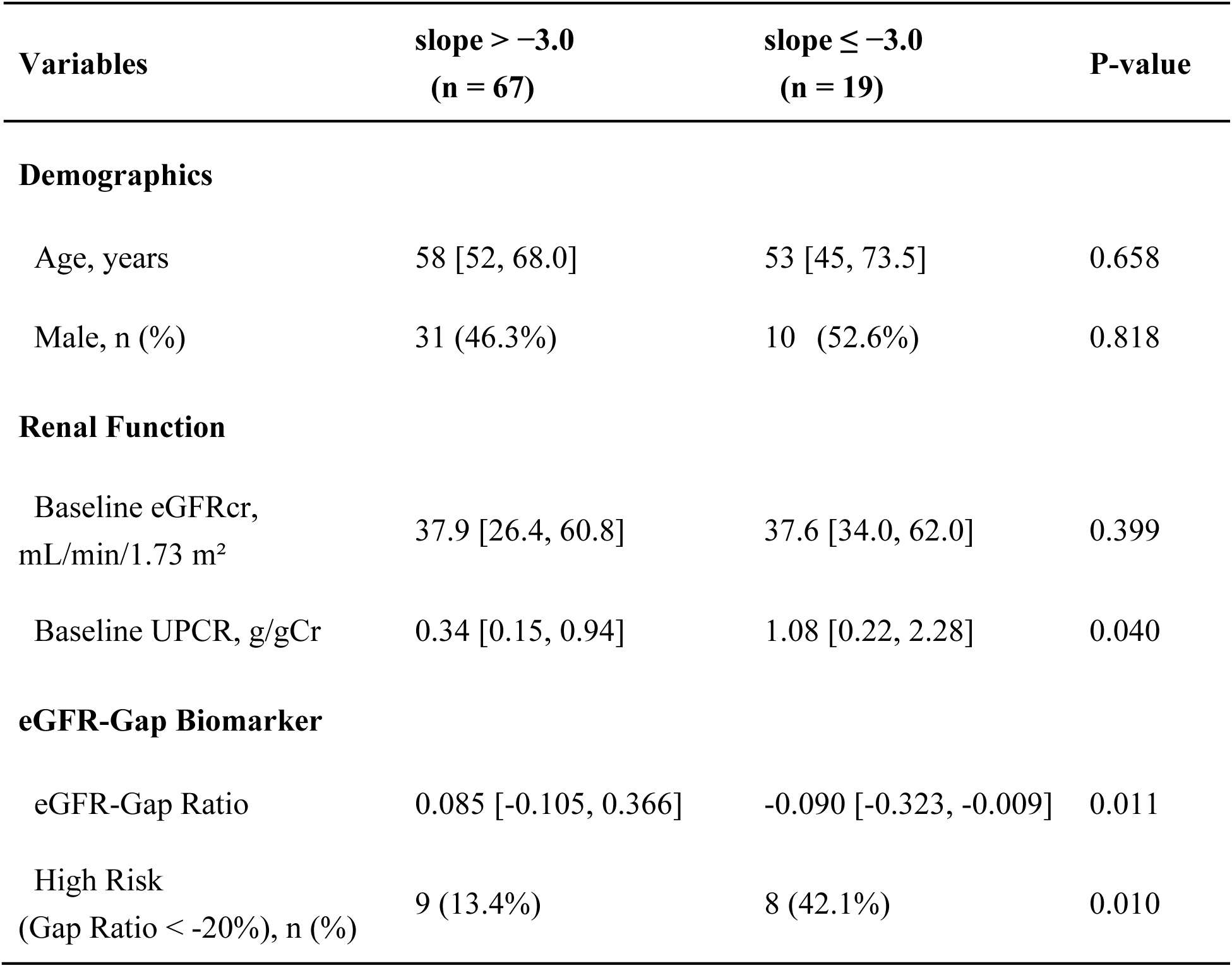
Baseline clinical characteristics of patients stratified by a moderate progression threshold (slope ≤ −3.0 mL/min/1.73 m²/year). Moderate progression was defined as an annualized eGFRcr slope of ≤ −3.0 mL/min/1.73 m²/year from the date of urinary MUC1_CD9_ measurement. Continuous variables are expressed as mean ± s.d. or median (IQR) and categorical variables as n (%). P values were calculated using Student’s t-test, Mann–Whitney U test, or chi-square test, as appropriate. eGFRcr, creatinine-based estimated glomerular filtration rate; IQR, interquartile range; UPCR, urinary protein-to-creatinine ratio.

## References

1. Francis, A., et al. Chronic kidney disease and the global public health agenda: an international consensus. Nat Rev Nephrol 20, 473–485 (2024).

2. Lees, J.S., et al. Advances in the diagnosis and detection of chronic kidney disease. Lancet (2026).

3. Ostrominski, J.W., et al. Cardiovascular, kidney, and metabolic health: an actionable vision for heart failure prevention. Lancet 406, 1171–1192 (2025).

4. Fernández-Fernandez, B., Sarafidis, P., Soler, M.J. & Ortiz, A. EMPA-KIDNEY: expanding the range of kidney protection by SGLT2 inhibitors. Clin Kidney J 16, 1187–1198 (2023).

5. Heerspink, H.J.L., et al. Dapagliflozin in Patients with Chronic Kidney Disease. N Engl J Med 383, 1436–1446 (2020).

6. Herrington, W.G., et al. Empagliflozin in Patients with Chronic Kidney Disease. N Engl J Med 388, 117–127 (2023).

7. Perkovic, V., et al. Canagliflozin and Renal Outcomes in Type 2 Diabetes and Nephropathy. N Engl J Med 380, 2295–2306 (2019).

8. Group, K.D.I.G.O.K.C.W. KDIGO 2024 Clinical Practice Guideline for the Evaluation and Management of Chronic Kidney Disease. Kidney Int 105, S117–S314 (2024).

9. Kushner, P., Khunti, K., Cebrián, A. & Deed, G. Early Identification and Management of Chronic Kidney Disease: A Narrative Review of the Crucial Role of Primary Care Practitioners. Adv Ther 41, 3757–3770 (2024).

10. Ortiz, A., Yanagita, M., Yokoi, H. & Torra, R. Evolving strategies for early diagnosis, proactive prevention and treatment of CKD. Nephrol Dial Transplant (2025).

11. Mashayekhi, M., et al. Is It Time to Consider Population-Based Urine Dipstick Screening for Early Detection of Kidney Disease? Kidney Int Rep 11, 1–5 (2026).

12. Ruilope, L.M., et al. Prevention of cardiorenal damage: importance of albuminuria. Eur Heart J 44, 1112–1123 (2023).

13. Ortiz, A., et al. Preventing chronic kidney disease and maintaining kidney health: conclusions from a Kidney Disease: Improving Global Outcomes (KDIGO) Controversies Conference. Kidney Int 108, 555–571 (2025).

14. Kusunoki, H., et al. Relationships between cystatin C- and creatinine-based eGFR in Japanese rural community-dwelling older adults with sarcopenia. Clin Exp Nephrol 25, 231–239 (2021).

15. Rahn, K.H., Heidenreich, S. & Brückner, D. How to assess glomerular function and damage in humans. J Hypertens 17, 309–317 (1999).

16. Estrella, M.M., et al. Discordance in Creatinine- and Cystatin C-Based eGFR and Clinical Outcomes: A Meta-Analysis. JAMA 334, 1915–1926 (2025).

17. Tanaka, T., et al. Population characteristics and diagnosis rate of chronic kidney disease by eGFR and proteinuria in Japanese clinical practice: an observational database study. Sci Rep 14, 5172 (2024).

18. Freedman, B.I. & Cohen, A.H. Hypertension-attributed nephropathy: what’s in a name? Nat Rev Nephrol 12, 27–36 (2016).

19. Stonebrook, E., Hoff, M. & Spencer, J.D. Congenital Anomalies of the Kidney and Urinary Tract: A Clinical Review. Curr Treat Options Pediatr 5, 223–235 (2019).

20. Porrini, E., et al. Non-proteinuric pathways in loss of renal function in patients with type 2 diabetes. Lancet Diabetes Endocrinol 3, 382–391 (2015).

21. van Royen, M.E., et al. The quick reference card “Storage of urinary EVs” - A practical guideline tool for research and clinical laboratories. J Extracell Vesicles 12, e12286 (2023).

22. Grange, C. & Bussolati, B. Extracellular vesicles in kidney disease. Nat Rev Nephrol 18, 499–513 (2022).

23. Erdbrügger, U., Hoorn, E.J., Le, T.H., Blijdorp, C.J. & Burger, D. Extracellular Vesicles in Kidney Diseases: Moving Forward. Kidney 360 4, 245–257 (2023).

24. Rudolphi, C.F., Blijdorp, C.J., van Willigenburg, H., Salih, M. & Hoorn, E.J. Urinary extracellular vesicles and tubular transport. Nephrol Dial Transplant 38, 1583–1590 (2023).

25. Blijdorp, C.J., et al. Nephron mass determines the excretion rate of urinary extracellular vesicles. J Extracell Vesicles 11, e12181 (2022).

26. Blijdorp, C.J., Burger, D., Llorente, A., Martens-Uzunova, E.S. & Erdbrügger, U. Extracellular Vesicles as Novel Players in Kidney Disease. J Am Soc Nephrol 33, 467–471 (2022).

27. Takizawa, K., et al. Urinary extracellular vesicles signature for diagnosis of kidney disease. iScience 25, 105416 (2022).

28. Harita, Y. Urinary extracellular vesicles in childhood kidney diseases. Pediatr Nephrol 39, 2293–2300 (2024).

29. Bolignano, D., et al. Neutrophil gelatinase-associated lipocalin (NGAL) and progression of chronic kidney disease. Clin J Am Soc Nephrol 4, 337–344 (2009).

30. Coppolino, G., et al. Urinary Neutrophil Gelatinase-Associated Lipocalin (NGAL) Predicts Renal Function Decline in Patients With Glomerular Diseases. Front Cell Dev Biol 8, 336 (2020).

31. Woo, K.S., et al. Urinary neutrophil gelatinase-associated lipocalin levels in comparison with glomerular filtration rate for evaluation of renal function in patients with diabetic chronic kidney disease. Diabetes Metab J 36, 307–313 (2012).

32. Levey, A.S., et al. Expressing the Modification of Diet in Renal Disease Study equation for estimating glomerular filtration rate with standardized serum creatinine values. Clin Chem 53, 766–772 (2007).

33. Levey, A.S., et al. A more accurate method to estimate glomerular filtration rate from serum creatinine: a new prediction equation. Modification of Diet in Renal Disease Study Group. Ann Intern Med 130, 461–470 (1999).

34. Cusick, M.M., et al. When to Start Population-Wide Screening for Chronic Kidney Disease: A Cost-Effectiveness Analysis. JAMA Health Forum 5, e243892 (2024).

35. Astley, M.E., et al. Age- and sex-specific reference values of estimated glomerular filtration rate for European adults. Kidney Int 107, 1076–1087 (2025).

36. Lake, B.B., et al. An atlas of healthy and injured cell states and niches in the human kidney. Nature 619, 585–594 (2023).

37. Project, T.K.P.M. The results here are in whole or part based upon data generated by the Kidney Precision Medicine Project. Accessed Dec. 23, 2025. (https://www.kpmp.org).

38. Lake, B.B., et al. Cellular and Spatial Drivers of Unresolved Injury and Functional Decline in the Human Kidney. bioRxiv (2025).

39. Blijdorp, C.J., et al. Comparing Approaches to Normalize, Quantify, and Characterize Urinary Extracellular Vesicles. J Am Soc Nephrol 32, 1210–1226 (2021).

40. Welsh, J.A., et al. Minimal information for studies of extracellular vesicles (MISEV2023): From basic to advanced approaches. J Extracell Vesicles 13, e12404 (2024).

41. Vanichapol, T., et al. Permanent defects in renal medullary structure and function after reversal of urinary obstruction. JCI Insight 10(2025).

42. Mukamel, R.E., et al. Protein-coding repeat polymorphisms strongly shape diverse human phenotypes. Science 373, 1499–1505 (2021).

43. Kirby, A., et al. Mutations causing medullary cystic kidney disease type 1 lie in a large VNTR in MUC1 missed by massively parallel sequencing. Nat Genet 45, 299–303 (2013).

44. Kuriyama, S., et al. The Tohoku Medical Megabank Project: Design and Mission. J Epidemiol 26, 493–511 (2016).

45. Kuriyama, S., et al. Cohort Profile: Tohoku Medical Megabank Project Birth and Three-Generation Cohort Study (TMM BirThree Cohort Study): rationale, progress and perspective. Int J Epidemiol 49, 18–19m (2020).

46. Hozawa, A., et al. Study Profile of the Tohoku Medical Megabank Community-Based Cohort Study. J Epidemiol 31, 65–76 (2021).

47. Takizawa, K., Nishimura, T. & Harita, Y. Enzyme-linked immunosorbent assay to detect surface marker proteins of extracellular vesicles purified from human urine. STAR Protoc 4, 102415 (2023).

48. Erdbrügger, U., et al. Urinary extracellular vesicles: A position paper by the Urine Task Force of the International Society for Extracellular Vesicles. J Extracell Vesicles 10, e12093 (2021).

49. Matsuo, S., et al. Revised equations for estimated GFR from serum creatinine in Japan. Am J Kidney Dis 53, 982–992 (2009).

50. Uemura, O., et al. Creatinine-based equation to estimate the glomerular filtration rate in Japanese children and adolescents with chronic kidney disease. Clin Exp Nephrol 18, 626–633 (2014).

51. Horio, M., et al. GFR estimation using standardized serum cystatin C in Japan. Am J Kidney Dis 61, 197–203 (2013).

52. Hanisch, F.G. & Ninkovic, T. Immunology of O-glycosylated proteins: approaches to the design of a MUC1 glycopeptide-based tumor vaccine. Curr Protein Pept Sci 7, 307–315 (2006).

53. Yoshimura, Y., et al. Products of Chemoenzymatic Synthesis Representing MUC1 Tandem Repeat Unit with T-, ST- or STn-antigen Revealed Distinct Specificities of Anti-MUC1 Antibodies. Sci Rep 9, 16641 (2019).

54. Perez, G.I., et al. Phosphatidylserine-Exposing Annexin A1-Positive Extracellular Vesicles: Potential Cancer Biomarkers. Vaccines (Basel) 11(2023).

55. Wettersten, N., et al. Urinary Biomarkers and Kidney Outcomes: Impact of Indexing Versus Adjusting for Urinary Creatinine. Kidney Med 3, 546–554.e541 (2021).

56. Aydoğdu, M., Oral, S. & Akgür, S.A. The impact of creatinine reference value: Normalization of urinary drug concentrations. J Forensic Sci 66, 1855–1861 (2021).

