## Supplemental Tables and Figures for "A non-invasive liquid biopsy resolves the diagnostic blind spot in chronic kidney disease"

**Contents:**

**Supplemental Table 1 |** Diagnostic performance of eGFRu at eGFRcr thresholds of < 45 and < 60 mL/min/1.73 m² across cohorts.
**Supplemental Table 2 |** Test–retest reliability of eGFRu and eGFRcr across consecutive outpatient visits.
**Supplemental Table 3 |** Clinical characteristics of the high-risk and low-risk groups stratified by Tim4-based eGFR-Gap Ratio.
**Supplemental Table 4 |** Impact of different Tim4 Gap Ratio thresholds on longitudinal eGFRcr decline: A Linear Mixed-effects (LME) model analysis.
**Supplemental Table 5 |** Baseline clinical characteristics of rapid and non-rapid progressors defined by Tim4-based eGFRu.
**Supplemental Table 6 |** Multivariable logistic regression models for predictors of rapid renal progression using High-Risk categorization based on Tim4 eGFR-Gap.

**Supplemental Figure 1 |** Development and validation of the Tim4-based eGFR model.
**Supplemental Figure 2 |** Longitudinal eGFRcr trajectories and predicted annual slopes stratified by Tim4 Gap Risk.
**Supplemental Figure 3 |** Performance Comparison of eGFR Estimation Models: Impact of Including Urinary Total Protein (uTP)

#### Supplemental Tables

**Supplemental Table 1 | Diagnostic performance of eGFRu at eGFRcr thresholds of < 45 and < 60 mL/min/1.73 $m^2$ across cohorts.**

| Cohort (Ref) | Population | Target | n | Pos | AUC | Sensitivity | Specificity | Accuracy |
|---|---|---|---|---|---|---|---|---|
| Discovery (eGFR$_{Cr}$) | Overall | < 45 | 471 | 109 | 0.897 | 0.789 | 0.865 | 0.847 |
| | | < 60 | 471 | 215 | 0.832 | 0.693 | 0.805 | 0.754 |
| | Age < 60 | < 45 | 183 | 37 | 0.967 | 0.892 | 0.932 | 0.923 |
| | | < 60 | 183 | 49 | 0.932 | 0.816 | 0.940 | 0.907 |
| Validation_HC (eGFR$_{Cr}$) | Overall | < 45 | 192 | 79 | 0.952 | 1.000 | 0.832 | 0.901 |
| | | < 60 | 192 | 98 | 0.935 | 0.908 | 0.904 | 0.906 |
| | Age < 60 | < 45 | 131 | 33 | 0.964 | 1.000 | 0.888 | 0.916 |
| | | < 60 | 131 | 47 | 0.937 | 0.957 | 0.821 | 0.870 |
| Validation_BC (eGFR$_{CrysC}$) | Overall | < 45 | 197 | 20 | 0.977 | 0.900 | 0.977 | 0.970 |
| | | < 60 | 197 | 38 | 0.925 | 0.921 | 0.786 | 0.807 |
| | Age < 60 | < 45 | 127 | 7 | 0.999 | 1.000 | 0.992 | 0.992 |
| | | < 60 | 127 | 15 | 0.971 | 0.867 | 0.973 | 0.961 |

Diagnostic metrics are shown for the overall population and for adults aged < 60 years. Sensitivity, specificity, and accuracy are expressed as proportions. AUC, area under the receiver operating characteristic curve; eGFRcr, creatinine-based estimated glomerular filtration rate; eGFR$_{CysC}$, cystatin C-based estimated glomerular filtration rate; n, total number of participants; Pos, number of positive cases.

**Supplemental Table 2 | Test–retest reliability of eGFRu and eGFRcr across consecutive outpatient visits.**

| **Parameter** | **Value** |
|---|---|
| Number of Pairs (N) | 192 |
| Mean Interval between Visits (Days) | 168.5 ± 127.6 |
| Pearson Correlation Coefficient (R) | 0.953 |
| Mean Bias (2nd - 1st Visit) | -0.78 |
| Standard Deviation of Bias | 8.89 |
| Mean Absolute Difference | 6.68 |

Data are from 192 pairs of consecutive outpatient visits; mean interval between measurements was 5.6 months. Mean bias, s.d. of bias, and mean absolute difference are expressed in mL/min/1.73 $m^2$. Mean interval is expressed as mean ± s.d. eGFRcr, creatinine-based estimated glomerular filtration rate; eGFRu, urinary MUC1-derived estimated glomerular filtration rate; n, number of pairs; R, Pearson correlation coefficient.

**Supplemental Table 3 | Clinical characteristics of the high-risk and low-risk groups stratified by Tim4-based eGFR-Gap Ratio.**

| Parameter | Overall (n = 86) | Low Risk (n = 77) | High Risk (n = 9) | P-value |
|---|---|---|---|---|
| **n** | 86 | 77 | 9 | - |
| **Age (years)** | 57.5 [50.0, 70.5] | 57.0 [50.0, 68.0] | 68.0 [46.0, 76.0] | 0.534 |
| **Female, n (%)** | 45 (52.3%) | 40 (51.9%) | 5 (55.6%) | 1.000 |
| **Baseline eGFRcr** | 37.9 [26.9, 61.1] | 37.9 [26.7, 59.0] | 64.6 [35.4, 83.5] | 0.096 |
| **Baseline eGFRu (Tim4)** | 50.0 [40.7, 54.8] | 50.1 [43.6, 54.7] | 35.0 [15.3, 55.0] | 0.085 |
| **Urinary TP/Cr (g/gCr)** | 0.38 [0.16, 1.23] | 0.34 [0.15, 1.17] | 1.05 [0.38, 2.50] | **0.041** |
| **Observation Period (yrs)** | 2.0 [1.5, 2.1] | 2.0 [1.7, 2.1] | 1.5 [1.4, 1.9] | 0.052 |
| **LME Annual Slope ($mL/min/1.73m^2/yr$)** | — | -0.90 | -4.51 | **0.004** |

Baseline characteristics and renal prognosis of the study population (n = 86) categorized by Tim4 Gap Risk. The High-Risk group (n=9) includes patients with a Gap Ratio <−0.30 (predicted eGFR is >30% lower than measured eGFRcr), while the Low-Risk group (n=77) includes all others.

Continuous variables are presented as median [interquartile range (IQR)], except for the LME Annual Slope, which is expressed as mean (standard error [SE]). Annual eGFRcr slopes and their standard errors (SE) were derived from the fixed-effect estimates of a linear mixed-effects model: eGFRcr∼Time×Group+(Time|ID). Categorical variables are presented as number (percentage).

**Supplemental Table 4 | Impact of different Tim4 Gap Ratio thresholds on longitudinal eGFRcr decline: A Linear Mixed-effects (LME) model analysis.**

| Gap Cutoff | High Risk (n) | Slope (Low Risk) | Additional Decline in High Risk | P-value (Interaction) |
|---|---|---|---|---|
| **-20%** | 15 | −0.978 | −1.747 | 0.0932 |
| **-30%** | 9 | −0.901 | −3.608 | **0.0043** |

This table compares the statistical performance and clinical relevance of two potential thresholds for the Tim4-based eGFR-Gap Ratio ($< -20\%$ and $< -30\%$).
LME Model Structure: A linear mixed-effects model was employed to account for the longitudinal nature of the data, incorporating individual random intercepts and random slopes. The model formula was: $eGFRcr \sim Time \times Group + (Time|ID)$.
High Risk (n): The number of patients identified as "High Risk" at each threshold.
Slope (Low Risk): The predicted annual eGFRcr decline rate (mL/min/1.73m 2 /year) for patients who did not meet the High-Risk criteria.
Additional Decline in High Risk: The interaction term (Time×Group) coefficient, representing the additional annual rate of eGFRcr decline observed in the High-Risk group compared to the Low-Risk group.
P-value (Interaction): The statistical significance of the difference in slopes between the two groups.

**Supplemental Table 5 | Baseline clinical characteristics of rapid and non-rapid progressors defined by Tim4-based eGFRu.**

| Parameter | Non-Rapid Progressor (n = 71) | Rapid Progressor (n = 15) | P_value |
|---|---|---|---|
| **Age, years** | 59.0 [51.5, 70.0] | 52.0 [45.0, 69.5] | 0.248 |
| **Male, n (%)** | 35 (49.3%) | 6 (40.0%) | 0.578 |
| **Baseline eGFRcr, mL/min/1.73 m2** | 37.9 [26.7, 59.5] | 48.1 [30.9, 71.4] | 0.292 |
| **Baseline UPCR, g/gCr** | 0.3 [0.2, 1.0] | 1.1 [0.2, 2.3] | 0.095 |
| **eGFR-Gap Ratio (Tim4)** | 0.199 [-0.092, 0.559] | -0.157 [-0.343, -0.027] | **0.002** |
| **High Risk (Gap Ratio (Tim4) < -20%), n (%)** | 8 (11.3%) | 7 (46.7%) | **0.004** |
| **High Risk (Gap Ratio (Tim4) < -30%), n (%)** | 4 (5.6%) | 5 (33.3%) | **0.007** |

Rapid progression was defined as an annualized eGFRcr slope of ≤−5.0 mL/min/1.73 m²/year from the date of urinary MUC1CD9 measurement. Continuous variables are expressed as mean ± s.d. or median (IQR) and categorical variables as n (%). P values were calculated using Student's t-test, Mann–Whitney U test, or chi-square test, as appropriate. eGFRcr, creatinine-based estimated glomerular filtration rate; IQR, interquartile range; UPCR, urinary protein-to-creatinine ratio.

The eGFR-Gap Ratio was calculated using the Tim4-based 643-formula. High Risk was defined using two thresholds: a Gap Ratio < −0.20 and a Gap Ratio < −0.30. The results demonstrate that while both thresholds are significant, the -30% threshold provides higher specificity (91.6%) for predicting rapid eGFR decline (Slope ≤ −5.0)."

**Supplemental Table 6 | Multivariable logistic regression models for predictors of rapid renal progression using High-Risk categorization based on Tim4 eGFR-Gap.**

| | Model 1 (Multivariable) | | Model 2 (UPCR adjusted) | |
|---|---|---|---|---|
| **Analysis 1: Tim4 Gap < -20%** | | | | |
| **High Risk (Gap < -20%)** | 4.89 (1.14-20.88) | **0.029** | 5.48 (1.42-21.07) | **0.012** |
| **Age, per year** | 1.00 (0.94-1.06) | 0.983 | - | - |
| **Sex (Female)** | 1.74 (0.46-7.24) | 0.420 | - | - |
| **Baseline eGFRcr, per unit** | 1.01 (0.97-1.04) | 0.664 | - | - |
| **Baseline UPCR, per g/gCr** | 1.42 (1.04-2.05) | **0.042** | 1.35 (1.02-1.88) | **0.048** |
| **Analysis 2: Tim4 Gap < -30%** | | | | |
| **High Risk (Gap < -30%)** | 5.98 (1.00-38.22) | **0.048** | 6.00 (1.18-30.49) | **0.026** |
| **Age, per year** | 0.99 (0.93-1.05) | 0.637 | - | - |

| | | | | |
|---|---|---|---|---|
| **Sex (Female)** | 1.68 (0.44-7.04) | 0.452 | - | - |
| **Baseline eGFRcr, per unit** | 1.01 (0.97-1.05) | 0.630 | - | - |
| **Baseline UPCR, per g/gCr** | 1.40 (1.01-2.02) | 0.051 | 1.35 (1.01-1.88) | 0.053 |

Logistic regression models were constructed to evaluate the independence of the Tim4-based eGFR-Gap as a predictor of rapid progression.

High Risk Definitions: Two thresholds for the Tim4 Gap Ratio were evaluated: $< -20\%$ and $< -30\%$.
Model 1: Adjusted for Age, Sex, Baseline eGFRcr, and Baseline Urinary Protein-to-Creatinine Ratio (UPCR).
Model 2: Adjusted for Baseline UPCR, the primary conventional clinical risk factor.

In both thresholds, the Tim4 High Risk status remained a significant and independent predictor of rapid renal function decline. Notably, the −30% threshold demonstrated the highest effect size (OR 6.00, 95% CI 1.18-30.49, $p = 0.026$), indicating a six-fold increase in the risk of rapid progression compared to the Low-Risk group, even after adjusting for proteinuria. These findings suggest that the Tim4 eGFR-Gap reflects tubular damage that is not fully captured by conventional clinical markers.

#### Supplemental Figure Legends

**Supplemental Figure 1 | Development and validation of the Tim4-based eGFR model.**

The eGFRu formula using Tim4 was derived from log-linear regression and is expressed as:

eGFRu(Tim4) =227.75 × $Age^{-0.2726}$ × $(MUC1_{Tim4}/uCre)^{0.2657}$

a, Scatter plot of eGFRu(Tim4) versus eGFRcr in the discovery cohort. b, Bland–Altman plot of agreement between eGFRu and eGFRcr. c, ROC curves of eGFRu for identifying stages of renal impairment defined by eGFRcr thresholds (< 30, 45, 60, and 90 mL/min/1.73 $m^2$). d–f, Subgroup analysis for patients aged < 60 years: scatter plot (d), Bland–Altman plot (e), and ROC curves (f) corresponding to panels a–c. Scatter plots show individual data points with linear regression lines and 95% confidence intervals (shaded areas). The correlation between eGFRcr and eGFRu was assessed using Pearson's correlation coefficient (a, d). ROC, receiver operating characteristic.

**Supplemental Figure 2 | Longitudinal eGFRcr trajectories and predicted annual slopes stratified by Tim4 Gap Risk.**

Spaghetti plots of individual longitudinal eGFRcr trajectories stratified by risk group. The High-Risk group (n = 9)(red) includes patients with a Gap Ratio < −0.30 (predicted eGFR is > 30% lower than measured eGFRcr), while the Low-Risk group (n = 77) (blue) includes all others.

The thin lines represent the observed individual trajectories of creatinine-based eGFR (eGFRcr) for each patient. The thick solid lines represent the population-level mean trajectories predicted by the linear mixed-effects (LME) model, accounting for individual random intercepts and slopes.

**Supplemental Figure 3 | Performance Comparison of eGFR Estimation Models: Impact of Including Urinary Total Protein (uTP)**

a, ROC curves of the eGFRu model (237.5 × $Age^{-0.278}$ × $(MUC1_{CD9}/uCre)^{0.256}$) for identifying stages of renal impairment (eGFRcr thresholds: < 30, 45, 60, and 90 mL/min/1.73 $m^2$) in the combined cohort (Discovery and Validation-HC, *n* = 433). b, ROC curves of the modified model including uTP (278.7 × $Age^{-0.358}$ × $uCre^{-0.198}$ × $MUC1_{CD9}{}^{0.238}$ × $uTP^{-0.042}$) at the same thresholds. Colour-coded lines represent different eGFR thresholds with corresponding AUC values. AUC, area under the receiver operating characteristic curve; ROC, receiver operating characteristic; uCre, urinary creatinine; uTP, urinary total protein (mg/dL).

### Supplemental Figure 1

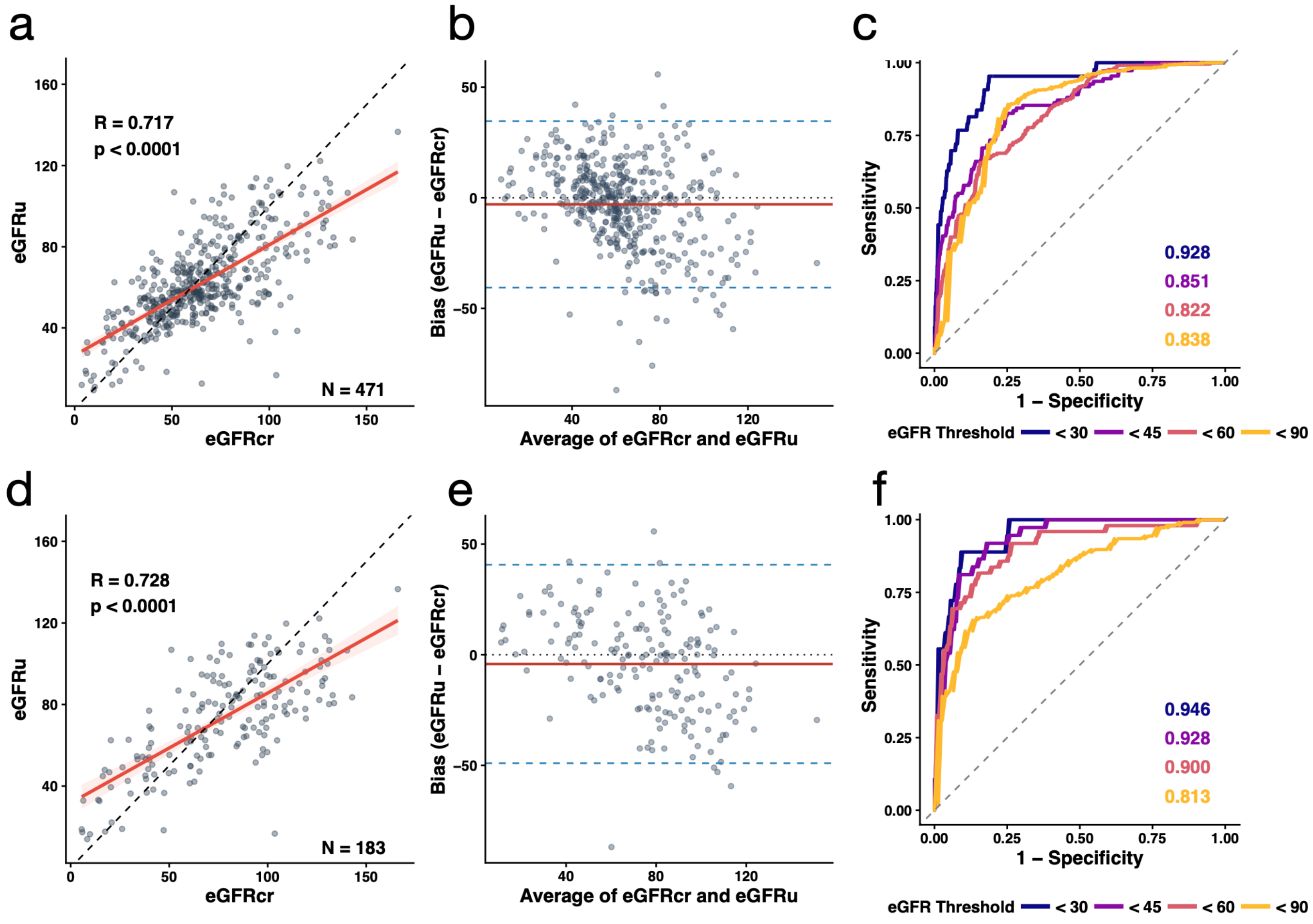

#### Supplemental Figure 2

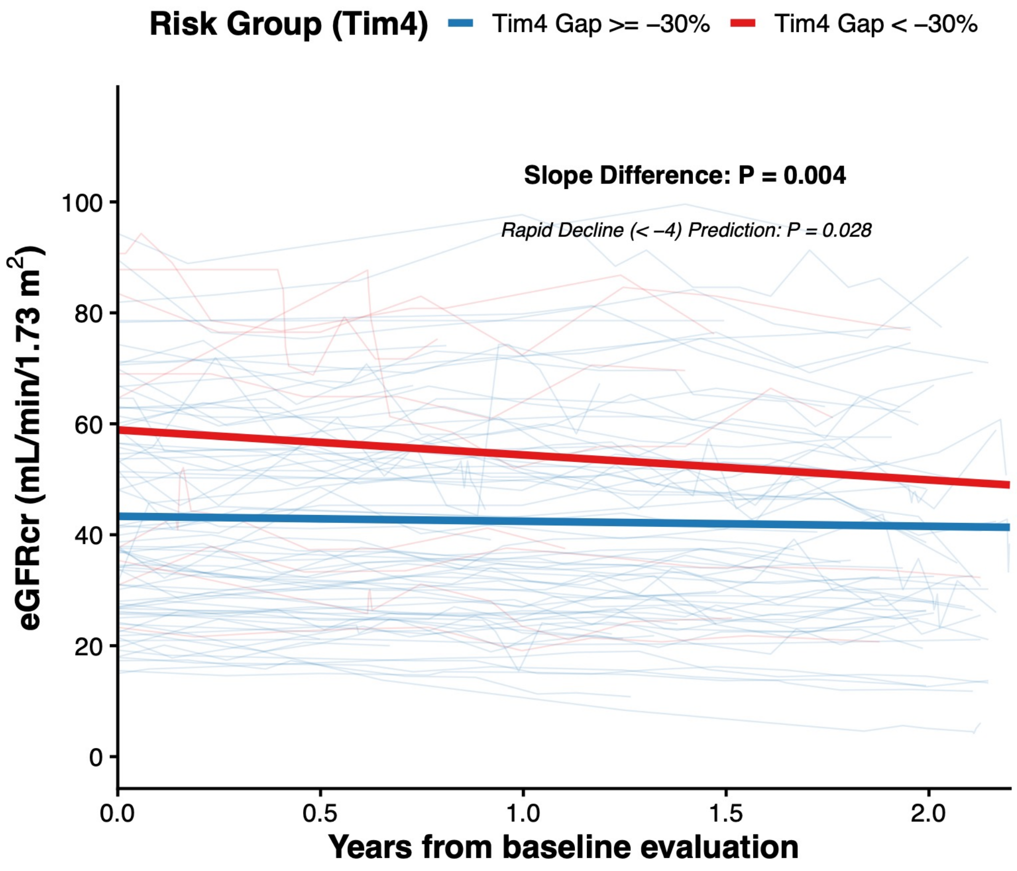

#### Supplemental Figure 3

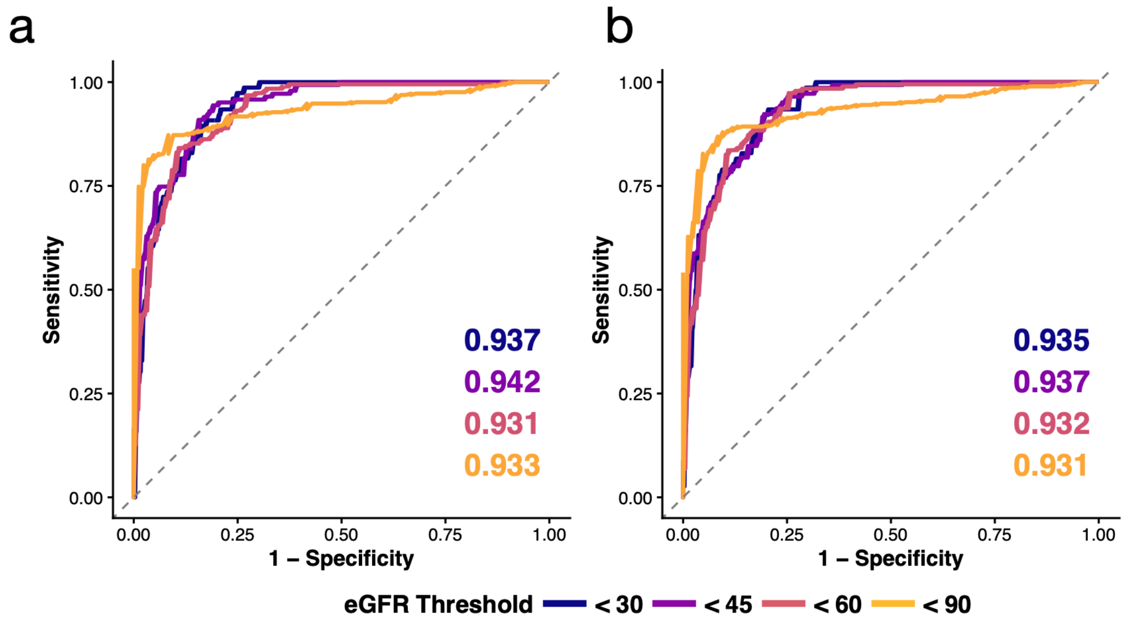
